# Quantitative Measurements of α-Synuclein Seeds in CSF Inform Diagnosis of Synucleinopathies

**DOI:** 10.1101/2025.03.06.25323462

**Authors:** Ilham Y. Abdi, Indulekha P. Sudhakaran, Simona S. Ghanem, Nishant N Vaikath, Nour Majbour, Yee Y. Goh, Nirosen Vijiaratnam, Christine Girges, Vasilios C Constantinides, Elisabeth Kapaki, George P. Paraskevas, Sandrina Weber, Gholam Adeli, Kostas Vekrellis, Daniel Erskine, Michele Hu, Thomas Foltynie, Henry Houlden, Laura Parkkinen, Wilma D.J. van de Berg, Brit Mollenhauer, Michael G. Schlossmacher, Omar M.A. El-Agnaf

**Affiliations:** Neurological Disorder Research Center, Qatar Biomedical Research Institute (QBRI), Hamad Bin Khalifa University (HBKU), Qatar Foundation; Doha, Qatar; College of Health and Life Sciences, Hamad Bin Khalifa University (HBKU), Qatar Foundation; Doha, Qatar; Neurochemistry and Biomarkers Unit, 1st Department of Neurology, National and Kapodistrian University of Athens; Athens, Greece; Ward of Cognitive and movement Disorders, 1st Department of Neurology, National and Kapodistrian University of Athens; Athens, Greece; Department of Neurology, University Medical Center Göttingen; Göttingen, Germany; Paracelsus-Elena-Klinik; Kassel, Germany; Hamad Medical Corporation; Doha, Qatar; Biomedical Research Foundation of the Academy of Athens; Athens, Greece; Translational and Clinical Research Institute, Newcastle University; Newcastle, United Kingdom; Nuffield Department of Clinical Neurosciences, Oxford Parkinson’s Disease Centre, University of Oxford; Oxford, UK; Amsterdam UMC Vrije University Amsterdam, Department of Anatomy and Neurosciences; Amsterdam, Netherlands; Neuroscience Program, The Ottawa Hospital; Ottawa, Ontario, Canada; University of Ottawa Brain and Mind Research Institute; Ottawa, Ontario, Canada; Division of Neurology, The Ottawa Hospital; Ottawa, Ontario, Canada; Translational Medicine, Neuroscience, Pharmaceuticals R&D, AstraZeneca; Cambridge, UK; Neuromuscular Diseases, UCL Queen Square Institute of Neurology, London, UK; Department of Clinical and Movement Neurosciences, UCL Queen Square Institute of Neurology, London, UK; Movement Disorders Centre, UCL Queen Square Institute of Neurology, London, UK

**Keywords:** α-synuclein, synucleinopathies, biomarkers, immunoassays, SAA

## Abstract

Diagnosing α-synucleinopathies and assessing target engagement in trials is hindered by the lack of reliable biomarkers. Here, we introduce a first-in-kind quantitative, highly sensitive, and disease-specific diagnostic assay, named Seeding Amplification ImmunoAssay (SAIA), developed and validated to detect synucleinopathy-linked disorders. To this end, we used wide range of specimens, including 37 brain homogenates (BH) and 559 cerebrospinal fluid (CSF) samples from subjects with diverse synucleinopathy disorders, non-synucleinopathy diseases, idiopathic REM sleep behavior disorder (iRBD), and controls. SAIA generated robust amplification results detecting disease-related α-synuclein seeds in BH samples at attogram levels, as referenced to preformed fibrils of α-synuclein. Furthermore, we conducted side-by-side comparisons between SAIA and a traditional Seeding Amplification Assay (SAA), which revealed high concordance. Further, SAIA distinguished synucleinopathies from non-synucleinopathies and controls with sensitivities and specificities ranging between 80-100% and area under the curve values exceeding 0.9. SAIA also accurately identified 24/24 (100%) iRBD cases, considered a prodromal state of PD, with 100% sensitivity and 80% specificity. Further optimization of SAIA through timepoint analyses revealed that shorter incubation times enhanced the assay’s specificity for distinguishing MSA from PD highlighting the potential for improved differentiation between specific synucleinopathies. In conclusion, SAIA represents a novel powerful high throughput method to screen, diagnose and monitor synucleinopathy disorders in living subjects, offering significant improvements over existing methods to assist clinical trials that aim to test disease modifying interventions.

## Introduction

Synucleinopathies, including Parkinson’s disease (PD), dementia with Lewy bodies (DLB), and multiple system atrophy (MSA), are associated with the misfolding and aggregation of the protein α-synuclein into toxic insoluble fibrils in the selectively vulnerable regions of the brain.^1^ Despite the tangible progress made in our understanding of disease, there’s unmet need for establishing reliable and disease-specific biomarkers for diagnosis or progression of synucleinopathies.^2,3^ This need can only be achieved through developing innovative and robust methods and assays for biomarkers discovery and assessments.

Various approaches have been employed to detect misfolded α-synuclein aggregates in tissues and biological samples. Among them are antibody-based immunoassays like ELISA ^4–7^, as well as assays that exploit the self-propagating property of α-synuclein aggregates, such as seed amplification assay (SAA).^8,9^ Both approaches have demonstrated notable advantages in the detection and analysis of synucleinopathies in living subjects.^6,10–14^ Seeding assays, in particular, demonstrate high sensitivity and specificity in detecting α-synuclein aggregates at low levels and even at early stages of the disease, enabling early diagnosis.^15^ Several of these studies have shown remarkable specificity scores, effectively discriminating PD, DLB, MSA patients from non-synucleinopathies cases and enhancing our understanding of disease molecular diagnosis.^16–26^ On the other hand, ELISA platforms provide a versatile and widely used approach for quantifying α-synuclein protein levels and/or modified metabolites thereof.

Despite their effectiveness, both SAA and ELISA techniques have limitations. SAA despite its sensitivity and promising diagnostic potential lacks direct quantification capabilities, thus meaning it is labelled to a binary outcome of positive or negative, and lacking additional data to support stratification of cases for prognostics or clinical trials. Efforts made to correlate SAA outputs with clinical parameters involved SAA kinetics.^16,26–33^ Furthermore, these methods necessitate specialized technical expertise and well-equipped laboratories, making them less accessible in routine clinical settings ^34^. On the other hand, ELISAs, although widely used, have limited sensitivity in detecting low levels of pathological α-synuclein species in biological samples ^19,35–37^. Limitations in these techniques, albeit being the most advanced diagnostic techniques currently available, highlight a need for improvements that can allow for a significant advancement in clinical diagnostics of synucleinopathies.

With this in mind, we aimed to develop a novel immunoassay that not only amplified disease-associated α-synuclein seeds in samples but also quantified them within the same assay, which we’ve termed “seeding amplification immunoassay (SAIA)”. Herein we present proof-of-concept of the SAIA technique, its development as a high throughput platform, and our validation efforts regarding its application to diverse sample types, cohorts and distinct clinical conditions, thereby offering valuable insights into the diagnostic capabilities of the assay (Figure 1). The results underscore the assay’s effectiveness in detecting α-synuclein seeding activity in various biological matrices, that include 38 BH specimens and 559 CSF samples. Our findings highlight the versatile and diagnostic value of SAIA in different cohorts that include synucleinopathies and prodromal cases thereof.

**Figure 1.**

Overview of Study Design. Schematic presentation of study design. 1) Proof of concept was established in a small number of BH and CSF samples. 2) Assay was characterized by its sensitivity, robustness and validated in larger number of BH and CSF samples. 3) Assay was further validated in a larger comprehensive set of clinical well-described cohorts. 4) Explorative experiments to improve assay specificity were conducted by testing different sample incubation times (2, 4, 6, 8, and 18hrs) in BH samples followed by validation of the optimal incubation time (4hrs) in CSF samples.

## Materials and methods

### Experimental Design

Our experimental approach can be summarized in three main steps: 1.) “Seed Capture”- biofluid samples, containing α-synuclein seeds, are subjected to capture by the conformation-specific anti-α-synuclein antibody, 2.) “Seed Amplification”- simultaneously, the captured seeds are seeded by recombinant α-synuclein monomers present in a reaction mix promoted by optimal incubation and shaking conditions, and lastly 3.) “Seed Detection”- The amplified seeds are then detected with an anti-α-synuclein antibody, generating a signal that could be extrapolated to a standard curve to determine the concentration of amplified α-synuclein seeds in the samples (Figure 2a). This assay setup allows for the capture, amplification, and quantification of α-synuclein seeds exclusively in samples where these seeds were present. The conformation specific anti-α-synuclein Ab used for this study is our in-house produced and characterized proprietary monoclonal Ab 2A1 (QABY Biotech, Qatar)^38^.

**Figure 2.**
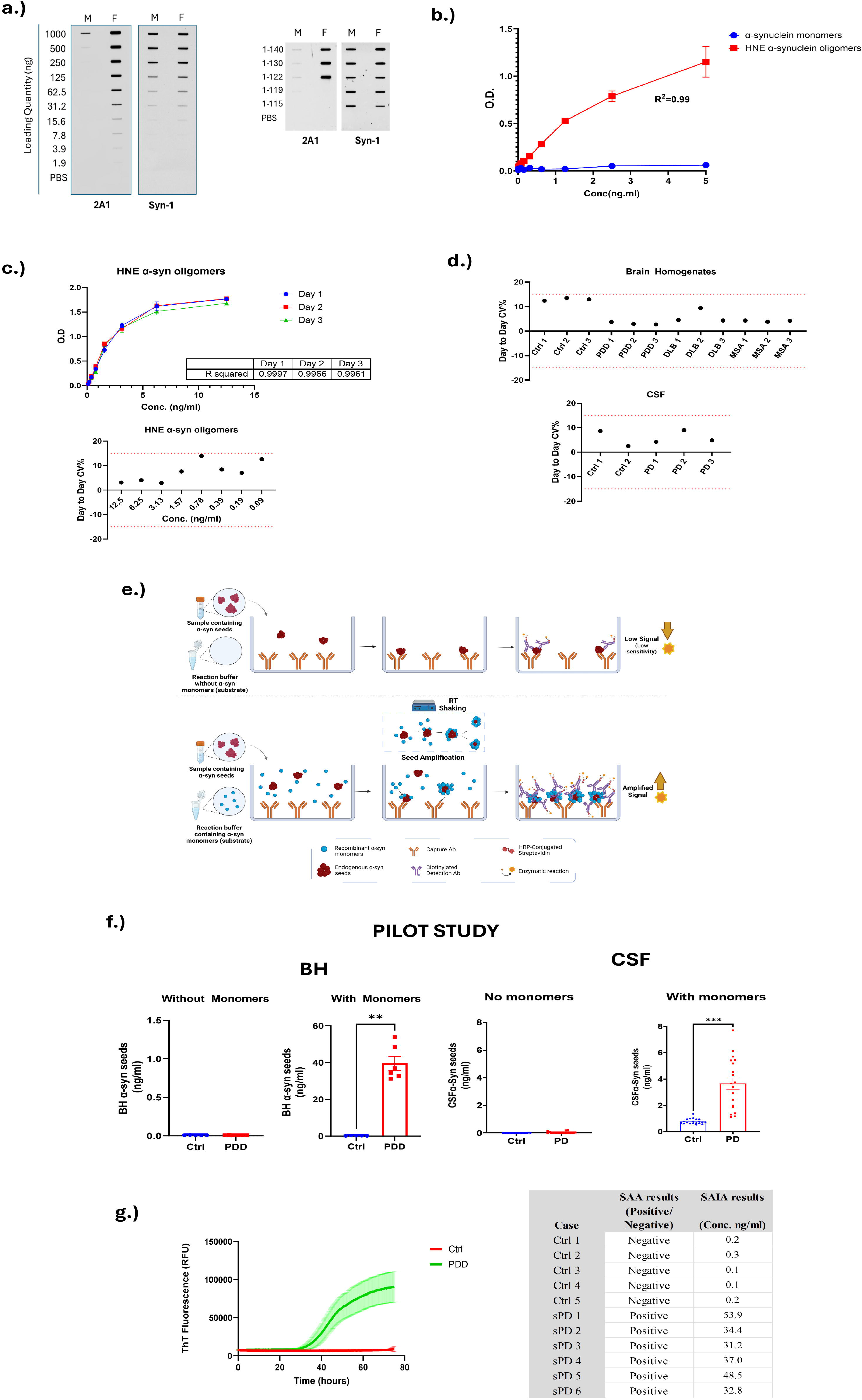
Illustration of seeding amplification immunoassay concept, assay characterization and proof of concept- Pilot study with BH and CSF. **a.)** Slot-blot assay showing our conformation-specific anti-α-synuclein mAb 2A1 preferentially detects α- synuclein aggregates, while Syn-1 binds both monomers and aggregates. 2A1 also recognizes full-length and truncated α-synuclein down to residue 122, whereas Syn-1 detects all fragments **b.) s**ELISA assay using 2A1 Ab showing selective detection of HNE-modified α- synuclein oligomers (red squares) compared to α-synuclein monomers (blue circles). A strong linear response (R² = 0.99) was observed for oligomers, while monomers showed minimal signal, confirming assay specificity for aggregated species. Data are presented as mean ± SD**. c.)** Inter-assay variability plots of HNE α-synuclein oligomer standards tested over three non-consecutive days. **d.)** BH and CSF samples tested over two non-consecutive days. **e.)** Diagrammatic representation of SAIA concept. The top panel of the diagram presents a conventional sandwich ELISA protocol employing without the seeding amplification step. In this setup, diluted samples are processed in a reaction buffer without adding recombinant α-synuclein monomer to serve as a substrate. Although this method allows for the capture and detection of endogenous α-syn seeds within the samples, it achieves this with limited sensitivity. The bottom panel illustrates an enhancement to this method under identical experimental conditions (same reaction buffer, shaking, and temperature). By introducing α-syn monomers, there is an amplification of endogenous α-syn seeds. This amplification could occur through two possible mechanisms: either directly in solution, leading to the formation of a complex aggregate of recombinant and endogenous alpha-synuclein captured by the monoclonal antibody, or by the initial capture of endogenous seeds by the antibody followed by seeding. Such amplification significantly increases the number of potential binding sites available for the detection antibody, biotinylated 2A1, thereby substantially enhancing the sensitivity and the overall signal detected in the assay. (Created with BioRender.com). f) Seeding effect, indicated by the increase in measured seeded α-synuclein, is evident in pilot study of PD BH and CSF samples respectively, in the presence of the substrate, α-synuclein monomers, whereas controls maintained a low signal. The error bars represent the standard error of the mean **g.)** SAA reactions of BH samples were performed in triplicate from both control individuals (red) and patients with PDD (green). The solid line denotes the mean ThT signal across triplicate wells, while the colored ribbon signifies the standard error.

The proof-of-concept was first conducted a pilot study using BH and CSF samples from individuals with PD and age-matched controls (n= 6/group for BH, n=20/group for CSF). All brain samples were taken from the frontal cortex, specifically the superior frontal gyrus. Results were validated through a side-by-side comparison with traditional SAA, with both assays confirming disease-specific α-synuclein seeding activity in PDD and PD samples.

The sensitivity of the assay as explored by testing serial dilutions, 1:100 to 1:204.2k, of BH from PD, DLB, MSA to assess the lowest dilution that can be used to amplified and quantified. Similarly, we also tested spiking dilutions of α-synuclein pre-formed fibrils in simple buffer and BH from control cases for a comparison between the assay’s sensitivity to endogenous α-synuclein compared to recombinant pre-formed fibrils.

A larger validation cohort of brain homogenate samples from subjects with PDD (n=10), DLB (n=10), and MSA (n=8), and controls (n=10). We then validated SAIA using CSF cohorts from the UK (Oxford Discovery; PD n=60 and Ctrl n=15) and the Netherlands (Progress-PD; PD n=47 and Ctrl n=51). We conducted an SAA for the BH samples and used previously published SAA results ^26^ of the UK (Oxford Discovery) CSF cohort to compare assay performances between SAA and SAIA. This phase evaluated the assay’s diagnostic sensitivity and specificity for PD.

The assay was further evaluated for its performance in CSF samples by validating in other CSF cohorts. The cohorts covered various disease groups to evaluate its 1.) Sensitivity at detecting at risk prodromal cases of synucleinopathies- German (DeNoPa) cohort (RBD vs Ctrl, n= 19 and 51 respectively) and, UK (Oxford Discovery) cohort (RBD n=5 vs Ctrl n=15) 2.) specificity to synucleinopathies compared to non-synucleinopathies-Greek cohort (PD, PDD, DLB, and MSA patients (n=34,12, 26, and 36, respectively), as well as non-synucleinopathy including progressive supranuclear palsy (PSP), Alzheimer’s disease (AD), vascular dementia (VD), and mild cognitive impairment (MCI) patients (n=34, 46, 16, and 7, respectively).

To enhance SAIA specificity for differentiating between synucleinopathies like PD and MSA, timepoint experiments were conducted. BH samples from PD and MSA cases were incubated for varying durations, ranging from 2 to 18 hours, to investigate time-dependent aggregation kinetics. Following the BH analysis, CSF samples were tested at selected timepoints to assess the influence of incubation times on specificity. This iterative approach was designed to determine optimal incubation conditions for distinguishing PD and MSA based on aggregation behavior, aiming to refine SAIA’s diagnostic capabilities.

This comprehensive experimental design allowed us to rigorously evaluate the SAIA’s performance across a range of sample types and clinical conditions, providing valuable insights into its potential clinical utility in synucleinopathy research and diagnosis.

### Clinical Cohorts

#### Post-mortem human brain tissues (Newcastle Cohort)

Brain tissue was obtained from Newcastle Brain Tissue Resource, a UK Human Tissue Authority-approved research tissue repository, and ethical approval was granted by the Joint Ethics Committee of Newcastle and North Tyneside Health Authority (ref: 19/NE/0008).

At autopsy, tissue was prepared by removing the brainstem at the level of the red nucleus and the cerebrum hemisected through the corpus callosum. The right hemisphere was immersion-fixed in neutral buffered formalin for subsequent paraffin wax embedding for neuropathological diagnosis, based on standardized neuropathological scales 70. The left hemisphere was sliced into 1 cm sections and rapidly frozen at -120°C between copper plates for molecular analysis.

PDD (n=10) and DLB (n=10) cases were identified and thus included in the present study on the basis of clinico-pathological diagnosis of either PDD or DLB by review of clinical records and neuropathological post-mortem report by experienced senior Old Age Psychiatrists and Neuropathologists, and on the basis of established international guidelines 71. MSA cases were included on the basis of clinical impression of MSA (n=6 parkinsonian type (MSA-P) and two cerebellar type (MSA-C)) and the presence of α-synuclein-immunoreactive Papp-Lantos bodies/glial cytoplasmic inclusions on neuropathological post-mortem examination 72. Control cases were included on the basis of an absence of α- synuclein pathology and did not show any other prominent protein pathology.

### CSF Samples

#### Netherlands (Progress-PD) Cohort

CSF samples collected from 47 PD patients and 51 age-matched controls, recruited at the Amsterdam UMC outpatient clinic were included ^39,40^. The study was approved by the local medical ethical committee of VU University Medical Center, Amsterdam. All patients gave written informed consent at study entry for the use of clinical information and CSF material for scientific research purposes. All patients were diagnosed with PD by movement disorder specialists according to the United Kingdom Parkinson′s Disease Society Brain Bank clinical diagnostic criteria ^41^. The severity of the motor symptoms was assessed using the UPDRS-III.

#### Greek Cohort

All patients were examined from 2011 to 2020 at the ward of Cognitive and Movement Disorders of Eginition Hospital. A total of 212 patients were included for the purposes of this study, of whom 109 had synucleinopathy. These included 41 clinically established PD, six patients with probable PDD, 26 patients with probable DLB, and 36 patients with probable MSA. Additionally, 103 patients with a non-synucleinopathic underlying pathology were included, comprising 34 patients with PSP, 46 patients with probable AD, 16 patients with VaD, and 7 patients with MCI. Diagnoses were set based on established diagnostic criteria.^42–49^

#### German (DeNoPa) Cohort

In this single-center clinical cohort study, the DeNoPa cohort was utilized, including 51 matched HC and 19 individuals with video-supported polysomnographically (vPSG) verified idiopathic REM sleep behavior disorder (iRBD). Controls were frequency matched based on age, sex, and education, and were required to be between 40 and 85 years old, free from central nervous system conditions, and without a family history of idiopathic PD. The iRBD group was diagnosed clinically and confirmed through vPSG, with exclusion criteria for medication-induced iRBD and severe sleep apnea. Dopamine transporter imaging was performed in iRBD subjects, and iRBD diagnosis was based on video analysis of vPSG data using established criteria. Details of the in-and exclusion criteria for the cohort have been reported in previous publications.

#### UK (Oxford Discovery) Cohort

The UK cohort is made up of 60 PD patients, 5 iRBD patients and 15 controls from a large, longitudinal Discovery study at the Oxford Parkinson’s Disease Centre ^50^. PD patients were diagnosed using UK PD Brain Bank criteria and iRBD using polysomnographic evidence (ICSD3) by specialist neurologists. Ethical approval was secured from local committees, and all participants signed written informed consent in accordance with the Declaration of Helsinki.

### UK (UCL) Cohort

The cohort of CSF samples provided from the University College of London (UCL) is a made up of baseline CSF samples collected from MSA and PD patients in two studies: 13 MSA patients from the “Progressive Supranuclear Palsy-Corticobasal Syndrome-Multiple System Atrophy” (PROSPECT-M-UK) study^51^ and 30 MSA patients and 57 PD patients from the Exenatide Study^52^.

### Production and purification of α-synuclein

Full length human α-synuclein protein was expressed in E. coli BL21 (DE3) cells and purified as previously described ^38^. Post purification by gel filtration and ion exchange, the final protein fractions were pooled and dialyzed against 1X PBS. Dialyzed proteins were filtered in a 100-kDa MWCO filter (Merck Millipore) and the protein concentrations were quantified using the Pierce BCA protein assay kit (Thermo Fisher). Purified stock protein was stored at -80oC. For seeding amplification assays, the stock protein is thawed, refiltered and quantified prior to each experiment.

### Filter Retardation Assay for antibody characterization

Serial dilutions of full-length human α-synuclein monomers and aggregates (ranging from 1000 ng to 0 ng), along with truncated α-synuclein variants (1–107, 1–115, 1–119, 1–122, 1–123, 1–130, 1–135, and 1–140) at 50 ng, were applied to a nitrocellulose membrane using a filtration apparatus. The membrane was probed with 2A1 and Syn-1 antibodies (50 µg/mL; Syn-1 from BD Biosciences), followed by incubation with HRP-conjugated goat anti-mouse secondary antibody (1:20,000 dilution). Detection was performed using SuperSignal™ West Pico PLUS Chemiluminescent Substrate (Thermo Fisher Scientific).

### Preparation of brain homogenates

Frozen brain tissue from frontal cortex (Brodmann Area 9) was dissected and approximately 0.1 mg of brain tissue is weighed out and added to 900 µl of TBS buffer (50mM Tris-HCl, 175 mM NaCl, pH 7.4) with protease inhibitors (1X). Homogenization was done using the Precellys homogenization machine (Cryolys - Bertin) using their custom soft tissue homogenization CK14 tubes. Two cycles of 15 sec homogenization were done at 5000 rpm with 5 min waiting time in between. Homogenate was centrifuged at 14000g, at 4 C for 3 min. The supernatant from this was treated as the soluble TBS fraction used for the assays. The total protein concentration was measured for each sample using BCA protein assay kit (Pierce, Thermo Scientific). Samples were then diluted and aliquoted at a final concentration of 1mg/ml. Aliquots were stored at -80 C till use.

### Seeding amplification Immunoassay (SAIA) for brain homogenates and CSF

For the seeding amplification immunoassay, 384 well plates (Nunc MaxiSorp, NUNC) were coated with anti-α-synuclein mAb 2A1 as capture antibody (at 0.5 µg/ml for BH analysis and 1 µg/ml for CSF analysis) in 0.2M NaHCO3 and incubated overnight (ON) at 4 C. After washing plate 3x (with PBS containing 0.05% tween-20; PBST) the plate was blocked for 2hrs at 37 C (in PBST containing 2.5 % gelatin) followed by a second wash. The brain homogenates were then added at a final total protein concentration of 5 µg/ml in 40 mM Phosphate buffer with 180 mM NaCl and 0.00375% SDS containing full length recombinant α-synuclein monomers (10 µg/ml) (50µl/well). For analysis of CSF samples, CSF was diluted to 50% with equal volume of reaction buffer containing α-synuclein monomers (25µg/ml) (50µl/well). Following addition of samples in reaction buffer, plates are incubated at RT overnight(18hrs) or 4hrs, on plate shaker set to 450 rpm. After a third washing step, Biotinylated 2A1 was used as detection with 1 hr incubation at 37 C (at 0.1 µg/ml for BH analysis and 0.25 µg/ml for CSF analysis). Plates were then incubated in Streptavidin HRP (Jackson ImmunoResearch) at 1:5000 for 1 hr at 37 C. Plates were incubated with TMB substrate (Supersignal ELISA Femto, Pierce) in dark for 10 min. Reaction was stopped with 0.6N H2SO4 and the plate was read at 450 absorbance in the microplate reader (PerkinElmer).

### SAIA assay characterization: Test of assay sensitivity

For the determination of minimal dilution of BH that can be detected by SAIA, dilutions of PDD, DLB and MSA brain homogenates were prepared and tested starting from a total protein concentration of 20 µg/ml (1:100 of the stock samples) followed by twelve 2x serial dilutions to reach a final dilution of 1:204k of the brain homogenate. Immediately prior to loading the samples, the prepared dilutions are added to the reaction buffer containing 10 µg/ml monomers.

Similarly, we tested to evaluate the SAIA sensitivity using recombinant preformed fibrils (PFFs). From a starting concentration of 5 ng/ml, PFFs were serially diluted 12 times until a final concentration of 0.05 ag/ml, and each dilution was spiked into reaction buffer containing 10 µg/ml α-synuclein monomers and control brain homogenates at 5 µg/ml or only 10 µg/ml α-synuclein monomers in artificial CSF.

### Seeding amplification assay (SAA) for brain homogenates and CSF

For brain homogenates, 160 μl of reaction mix composed of 40 mM phosphate buffer (pH 8.0), 1 mM citrate, 0.1 mg/mL recombinant monomeric αSyn (filtered through a 100 kD MWCO filter immediately before use) and 100 μM ThT was distributed in each well in a 96- well black plate with clear bottom (Nunc; Thermo Fisher, Waltham, MA) at a final volume of 200 μl per well. Reactions were performed in triplicates. For each test, we loaded 40 μl of BH of 0.1 mg/mL total protein concentration. The plate was then sealed with sealing tape and incubated in Omega FLUOstar plate reader (BMG Labtech) at 37°C with intermittent shaking cycles: double orbital with a 1-minute shake (500 rpm) and 15 minutes rest throughout the indicated incubation time. The sample was considered positive if two or more of the replicates were positive, otherwise the sample was classified as negative.

For CSF, wells were preloaded with 6 silica beads (Sigma-Aldrich, St. Louis, MO), and 85 μl of a reaction mix prepared to give final reaction concentrations of 40 mM phosphate buffer (pH 8.0), 170 mM NaCl, 0.1 mg/mL recombinant monomeric αSyn (filtered through a 100 kD MWCO filter immediately before use), 100 μM ThT, and 0.0015% sodium dodecyl sulfate was distributed in each well. Then, 15 μl of CSF per sample was spiked in triplicates into corresponding wells. The plate was then sealed with a sealing tape and incubated in Omega FLUOstar plate reader (BMG Labtech) at 42°C with intermittent shaking cycles: double orbital with a 1-minute shake (500 rpm) and 1-minute rest throughout the indicated incubation time. For both protocols, ThT fluorescence readings were taken every 25 minutes with a bottom read using 450 ± 10 nm (excitation) and 480 ± 10 nm (emission) wavelengths. The sample was considered positive if 2 or more of the replicates were above the calculated threshold. The threshold was calculated as the average fluorescence for all samples within the first 10 hours of incubation and 3 times the SDs.

### Statistical Analysis

The data were deemed unsuitable for parametric analyses following tests of normality. Spearman’s rank-order correlation coefficients were employed to explore correlations within the study cohorts. To assess distinctions between two diagnostic groups, the Mann–Whitney U test was employed. Meanwhile, the Kruskal-Wallis Test was applied for making comparisons across multiple groups. ROC Curve analysis was conducted to evaluate diagnostic parameters of the SAIA. A common threshold was obtained for all CSF samples using Youden Index calculation. The was done by calculating for a common threshold in all cohorts that provided a Youden index > 70% which was calculated to be 1.7ng/ml. Data analysis and creation of the corresponding graphs were performed using GraphPad Prism 9 (GraphPad Software, Inc., San Diego, CA).

## Results

### Establishing Proof-of-Concept for SAIA Using Postmortem Human Brain and CSF Samples

In this study, we utilized the in-house antibody 2A1, which has been extensively characterized both here and in previous publications^13^. This mouse IgG2a antibody specifically detects α-synuclein aggregates, as demonstrated by the filter retardation assay (Figure 2a), with minimal reactivity toward monomeric forms. It also detects fibrils prepared from truncated α-synuclein variants down to residue 122 (Figure 2a). Epitope mapping confirmed its binding to the C-terminal region (amino acids 113–123), while isotyping analysis determined it to be an IgG2a subclass antibody (Supplementary figure 1).

A sandwich ELISA was developed using 2A1as the capture antibody and biotinylated-2A1 for detection, incorporating HNE-modified α-synuclein oligomers as the standard calibrator. The assay displayed high linearity (R² = 0.99; Figure 2b), a detailed protocol for HNE-α- synuclein oligomers preparation was recently published by our group^53^. Monomeric α- synuclein was also tested in parallel and showed negligible signal, confirming the assay’s specificity for α-synuclein oligomers (Figure 2b).

These results validate the assay’s reliability in detecting α-synuclein aggregates with high specificity. The robustness of the assay was assessed by conducting repeat measurements of BH and CSF samples, as well as the calibrator standards, on three nonconsecutive days. Inter- and intra-assay variability were both observed to be less than 15% (Figure 2c–d).

Following a series of optimization steps, we demonstrated proof-of-concept of the SAIA technique in a small pilot study of *postmortem* BH samples. The BHs were derived from superior frontal cortex of individuals with clinico-pathologically-confirmed PDD and age-matched controls. The BHs were incubated for 18hrs at RT with continuous mixing (450rpm) in a reaction buffer containing recombinant α-synuclein monomers and compared to those in reaction buffer without α-synuclein monomers (Figure 2e). Samples incubated with the substrate, α-synuclein monomers, showed higher levels of seeded α-synuclein in PDD samples compared to control samples which showed negligible levels. In contrast, no seeded α-synuclein was detected in samples incubated without the presence of monomers (Figure 2b). This finding confirms the amplifications of α-synuclein seeds present in the PDD samples, specifically promoted by α-synuclein monomers acting as a substrate for the seeding reaction, thereby proving the concept of the SAIA.

We then explored SAIA application in CSF samples collected from living subjects using a pilot cohort of 20 PD subjects and 20 age-matched controls. Similar to BHs tested, CSF samples were incubated with and without the presence of α-synuclein monomers substrate. Consistent with the results obtained from BH samples, we observed a significant increase in α-synuclein seeding activity specifically in PD CSF samples, as evidenced by markedly higher levels of seeded α-synuclein compared to controls, which exhibited minimal α- synuclein seeding (p<0.001) (Figure 2f).

To further validate the results generated by SAIA, we performed a side-by-side comparison with the traditional SAA method. The SAA assay confirmed that PDD (brain) were SAA test-positive, while the control samples were negative (Figures 2g). These consistent results between SAIA and SAA underscore the reliability and accuracy of our novel assay in detecting disease-specific α-synuclein seeding activity in different biological matrices.

### Assay Sensitivity and Robustness Evaluation of SAIA: Definition of Detection Limits

Having confirmed that the SAIA concept works, we aimed to evaluate the assay’s sensitivity and robustness. To assess the sensitivity of the assay and determine the minimum quantity of α-synuclein seeds that could be accurately amplified and quantified, we conducted comprehensive investigations using two distinct approaches. Firstly, we performed serial dilutions of BH derived from individuals with PDD, MSA, and DLB, spanning dilutions from 1:100 to 1:204,200. Subsequently, these diluted samples were incubated in a reaction buffer containing α-syn monomers as a substrate. The results encouragingly demonstrated the assay’s remarkable sensitivity as it detected seeded α-synuclein in BH diluted up to 6,400 times while being incubated only for a short period of time (18hrs) and with a very low concentration of the substrate (10 µg/ml) (Figure 3a), exhibiting a concentration-dependent pattern as evident in the decreasing levels of seeded α-synuclein in diluted samples. Secondly, we employed α-synuclein preformed fibrils (PFFs) in a series of quantities, ranging from 5 nanograms to 0.05 attograms, incubated for 18hrs with a reaction buffer containing α- synuclein monomers as a substrate. This incubation was performed using BH obtained from pathologically-confirmed controls or artificial CSF (ACSF). Remarkably, our assay successfully amplified and quantified as little as 0.5 attogram of α-synuclein PFFs in the ACSF buffer and 500 attograms in control BH samples (Figure 3b).

**Figure 3.**
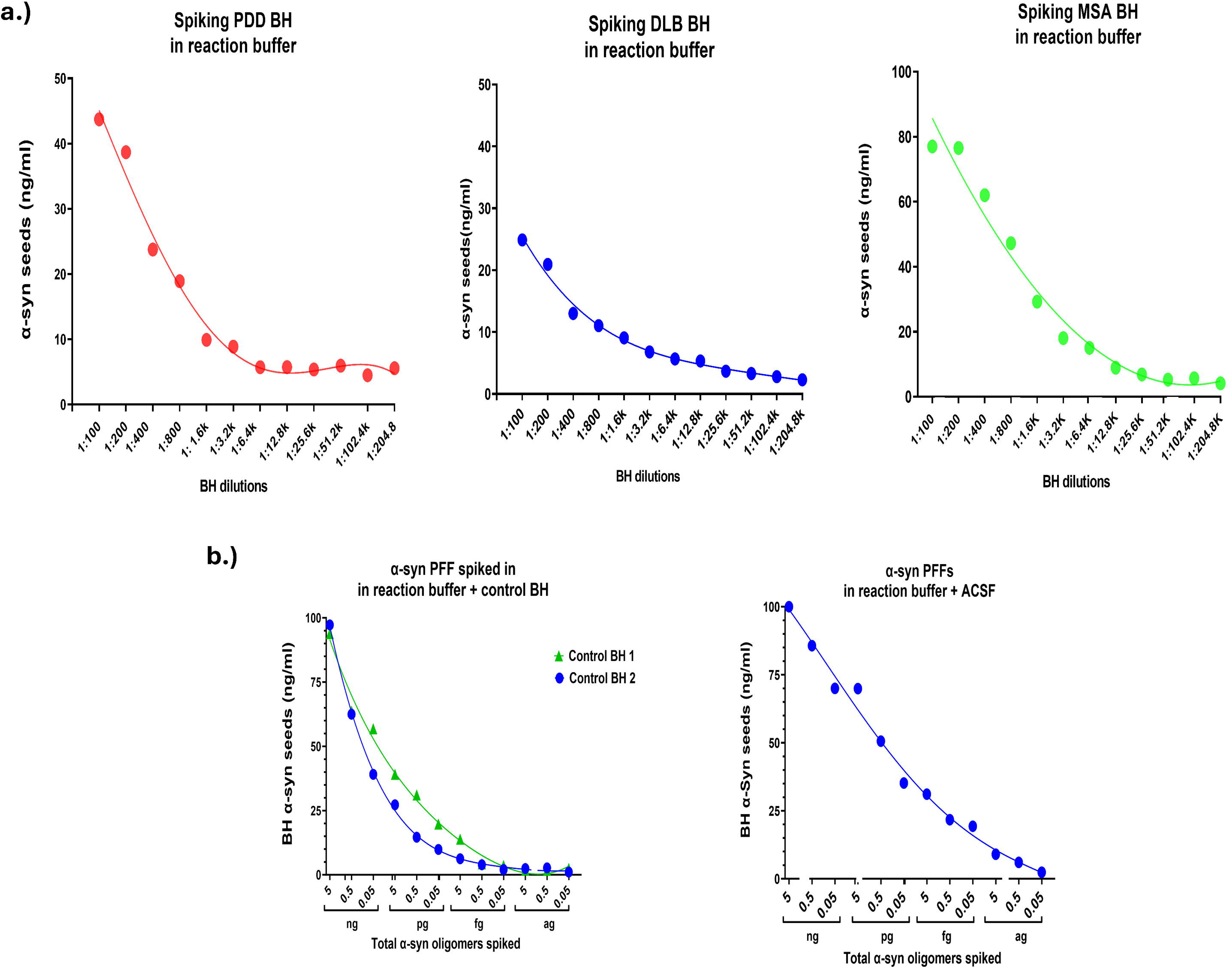
Characterisation of SAIA assay: sensitivity and robustness. **a)** The panel illustrates the SAIA assay’s sensitivity by determining the dilution, between 1:200 and 1:204.2k, of BH obtained from PD, DLB and MSA wherein seeded α-synuclein can be effectively detected. Sensitivity was established at approximate dilution of 1:6.4k of BH samples. **b)** SAIA’s detection capability is highlighted by assessing α-synuclein PFFs at series of dilutions from 5 nanograms to 0.05 attograms. Testing α-synuclein PFFs at different concentrations in control BH and artificial CSF (ACSF) showed sensitivity levels at 0.5 femtogram in BH and 0.5 attogram in ACSF buffer.

### Validating the SAIA Assay: Assessing CSF Seeded α-Synuclein Levels in Synucleinopathies and Controls

To validate the assay, we tested a larger cohort of *postmortem* BH samples, including those collected from subjects with clinic-pathologically confirmed PDD (n=10), DLB (n=10), MSA (n=6 MSA-P and 2 MSA-C), and controls (n=10); see Supplementary Table 1. We observed significantly higher levels of seeded α-synuclein in the synucleinopathies tested compared to controls. Using a threshold determined by ROC analysis to differentiate between positive and negative samples, we achieved a sensitivity of 100% and specificity of 90% for PDD, MSA, and 90% sensitivity and specificity for DLB samples against controls, with areas under the curve (AUC) values ranging from 0.99-1 (Figure 4a; Supplementary Table 2). We explored the correlation between the levels of seeded α-synuclein measured by SAIA and the neuropathological load of α-synuclein, quantified as the percentage area of LBs stained in six areas of interest within the same region (superior frontal gyrus) assessed by SAIA, but from the contralateral hemisphere in PDD and DLB cases. Unfortunately, brain sections from the same region were not available for the MSA cases. When assessing PDD and DLB cases together, no significant correlation was found between the mean percentage area of LB staining and the levels of seeded α-synuclein. However, when analyzed separately, a significant positive correlation was observed in PDD cases (r = 0.75, p < 0.05), suggesting a relationship between LB burden and α-synuclein seeding activity in this group. In contrast, no correlation was detected in DLB cases (Supplementary Fig 2).

**Figure 4.**
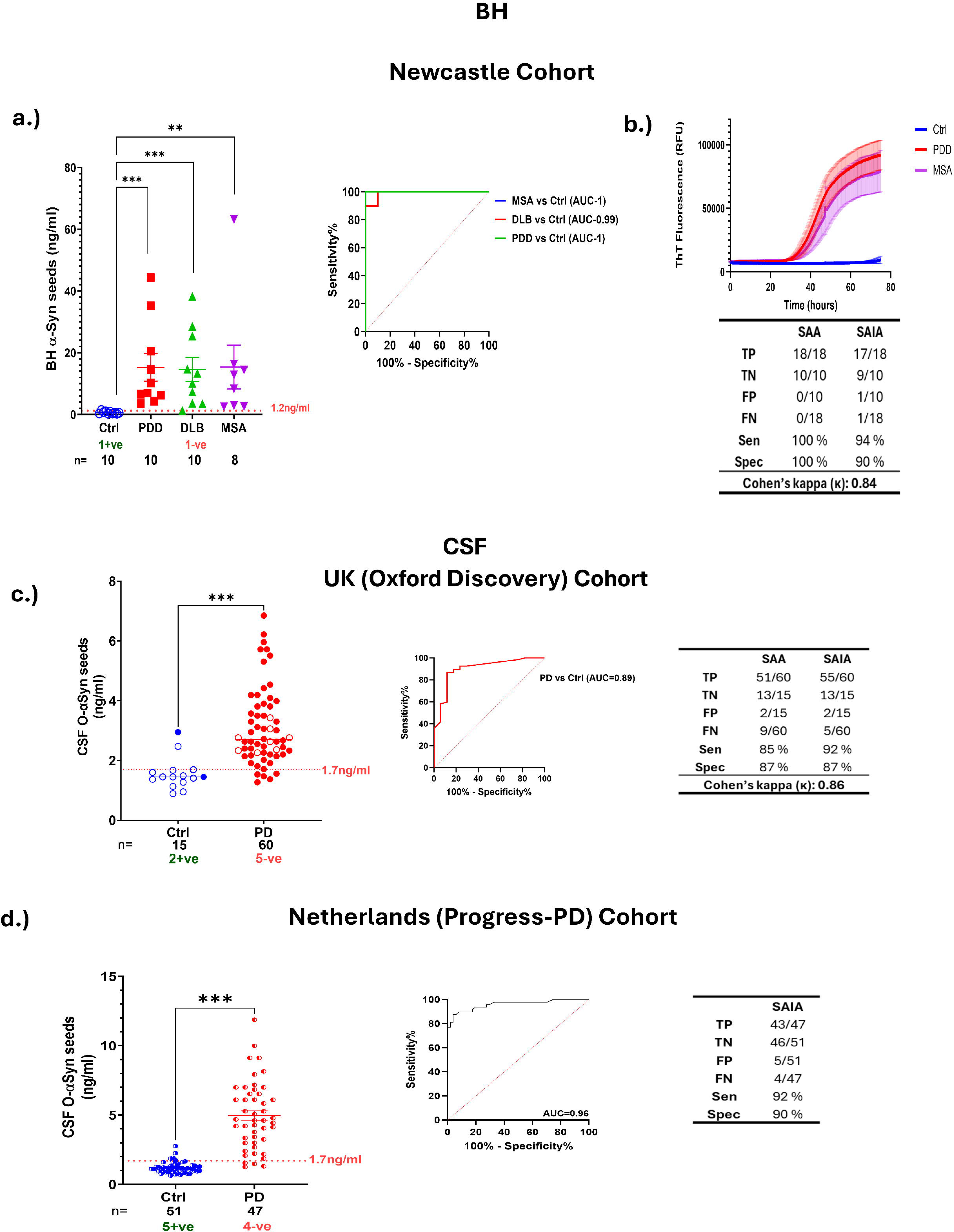
Validation of SAIA assay: BH and CSF analysis. **a.)** SAIA assay validated in a larger cohort of samples from brain homogenates from different synucleinopathies PDD, DLB, and MSA. Scatter plot and ROC analysis of measured seeded α-synuclein levels in BH samples of PDD, DLB, MSA vs control. **b.)** SAA reactions of BH samples seeded in triplicate from subjects confirmed with α-synuclein pathology (PDD, n = 10, MSA, n=8) compared to controls (n=9). The solid line represents the average ThT signal per group while error bars depict standard errors. **c, d.).** These panels show a scatter plot depicting the measured levels of seeded α-synuclein in CSF samples from two cohorts: Netherlands (Progress-PD) cohort (n= 51 controls and 47 PD), and UK (Oxford Discovery) cohort (n=15 controls and 60 PD). SAIA effectively distinguished PD cases from controls (p<0.001). CSF samples with signals above the threshold (1.7ng/ml red dashed horizontal line) calculated using Youden’s Index of ROC analyses were considered positive while those below were negative. This outcome underscores SAIA’s diagnostic potential as a robust tool for detecting PD. Open circles signify SAA-negative cases and closed colored circles illustrate SAA- positive cases. ROC curve analyses evaluate the diagnostic performance of SAIA in distinguishing between PD and control CSF samples and their corresponding sensitivities and specificities. *Statistical comparisons between groups were calculated by Kruskal-Wallis Test and Mann- Whitney U test for two group comparison (***- p<0.001, **-p<0.01). Cut-off of 1.2ng/ml for BH and 1.7ng/ml for CSF samples was calculated from Youden’s Index from ROC analysis. The error bars represent the standard error of the mean. Cohen’s kappa was calculated to measure the degree of agreement between SAA and SAIA*.

SAIA was validated for CSF samples by conducting a blind screening of two CSF cohorts: a Netherlands (Progress-PD) cohort (n=47 PD cases and 51 controls) and a UK (Oxford Discovery) cohort (n=60 PD cases and 15 controls). SAIA results indicated significantly higher levels of α-synuclein seeding activity in PD samples compared to controls (p<0.001) (Figure 4c-d; Supplementary Table 2). SAIA results of both cohorts showed clear distinction between PD and controls (p<0.001) with sensitivities and specificities above 90% (UK Oxford Discovery cohort: 13/15 negative controls and 55/60 positive PD cases ; Netherlands cohort: 46/51 negative controls and 43/47 positive PD cases).

To compare SAA and SAIA diagnostic performance, we tested the BH samples using SAA (Figure 4b) and used previously published SAA results for CSF samples from the UK (Oxford Discovery) cohort ^26^. The BH results showed all SAA positive cases except one DLB case to also be SAIA positive and similarly 9 out of the 10 control negative cases were negative in both assays. A total of 12 PD cases and 1 control of this UK cohort presented inconsistent results between SAA and SAIA. Of the 12 PD cases, 8 showed positive SAIA results but negative SAA results, while the other 4 showed the opposite. The degree of agreement between SAA and SAIA was assessed using Cohen’s kappa for both cohorts, yielding values of 0.84 for the BH cohort and 0.86 for the CSF cohort, indicating a high level of agreement. SAIA scatter plots in Figures 4 a and c illustrate SAA-positive samples as closed colored circles and SAA-negative samples as open circles. These samples were repeated in SAIA to confirm the results. The Netherlands cohort was not tested in SAA. These results demonstrated high concordance between SAIA and SAA, confirming the reliability of SAIA in detecting disease-specific α-synuclein seeding activity.

### Correlation Between SAA Kinetics and SAIA-Seeding Activity in BH and CSF Samples

We examined the relationship between SAA kinetics and SAIA-seeding activity across BH and CSF samples from the Newcastle and UK cohorts. Significant correlations were observed, demonstrating complementary findings between SAA and SAIA assays.

For BH samples from the Newcastle cohort, a strong negative correlation was found between T_50_ and SAIA seeded α-synuclein (r = -0.63, p < 0.001), indicating that faster aggregation times (shorter T50) correspond to higher seeded α-synuclein levels (Figure 5a). Positive correlations were observed between both AUC (r = 0.72, p < 0.001) and F_Max_ (r = 0.69, p < 0.001) with SAIA seeded α-synuclein, suggesting that higher seeding efficiency in SAA aligns with elevated α-synuclein levels detected by SAIA.

**Figure 5.**
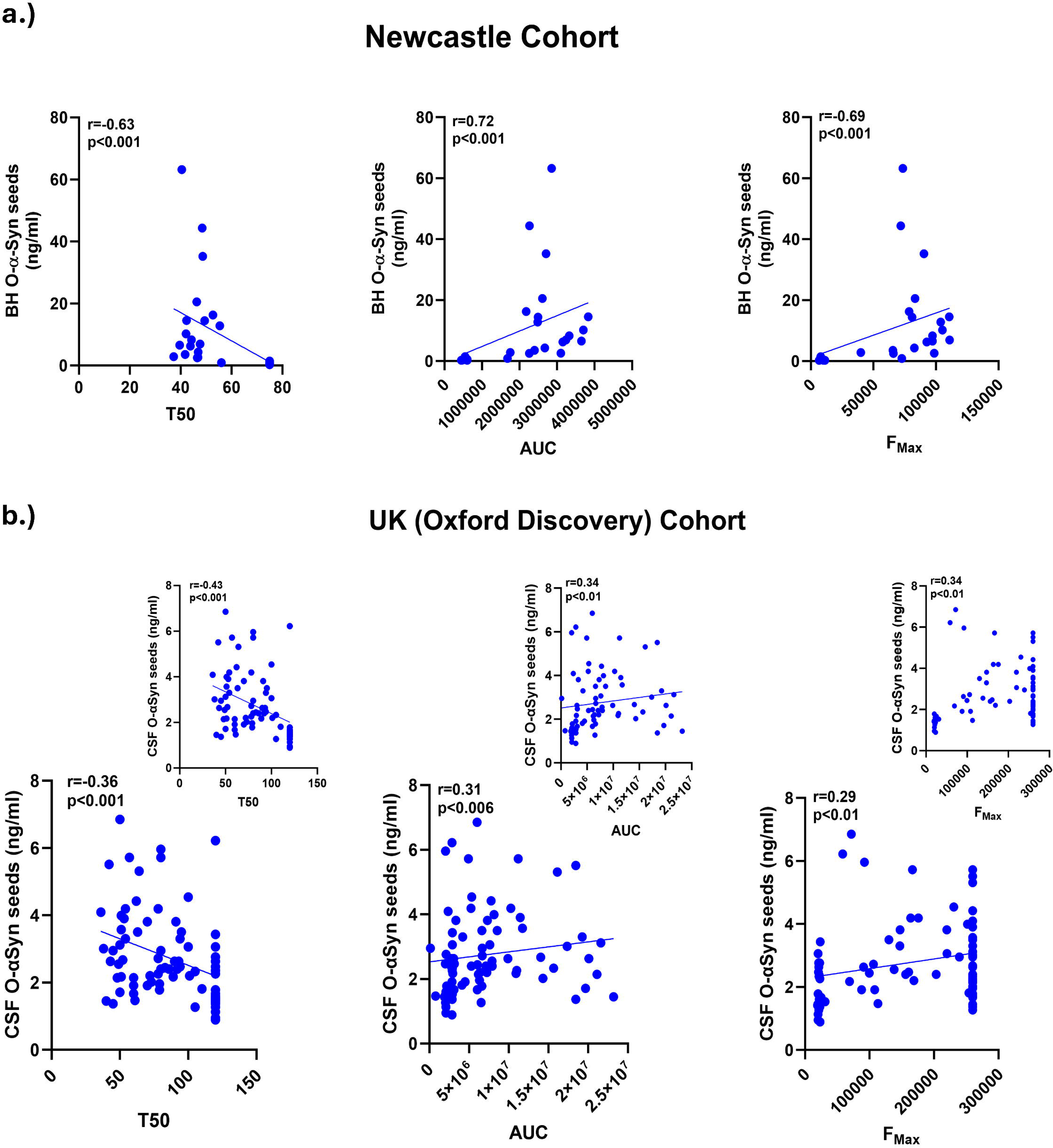
Correlation between SAA kinetics and SAIA-seeding activity in BH and CSF samples. a.) Scatter plots depicting correlations between SAA kinetic parameters (T_50_, AUC, F_Max_) and SAIA seeded α-synuclein levels in BH samples from the Newcastle cohort. T_50_ (left) shows an inverse relationship with SAIA seeded α-synuclein levels, while positive associations are observed with AUC (middle) and F_Max_ (right). b.) Scatter plots showing correlations between SAA kinetic parameters (T_50_, AUC, F_Max_) and SAIA seeded α-synuclein levels in cerebrospinal fluid (CSF) samples from the UK (Oxford Discovery) cohort. Similar trends are observed, with T_50_ (left) negatively associated with SAIA seeded α-synuclein levels, and positive associations with AUC (middle) and F_Max_ (right). Exclusion of discordant samples improves correlations, as indicated in the inset plots for each parameter. *Statistical analyses were performed using Spearman’s rank correlation coefficient to assess correlations between SAA kinetic parameters and SAIA seeded* α*-synuclein levels*.

In CSF samples, positive correlations were observed the correlations were notably weaker compared to BH samples (Figure 5b). There was a moderate negative correlation between T_50_ and SAIA-seeded α-synuclein levels (r = -0.36, p < 0.001), while a moderate positive correlation was observed between AUC or F_Max_ and SAIA seeded α-synuclein levels (AUC: r = 0.31, p = 0.006; F_Max_: r = 0.29, p < 0.01). Excluding the discordant samples between the SAA and SAIA improved the correlations observed to r=-0.43 for T_50_ and r=0.34 for AUC or FMax.

These findings collectively highlight the relationship between the kinetics of SAA and the quantitative measurements of α-synuclein seeding activity provided by SAIA, reinforcing the reliability and sensitivity of SAIA as a diagnostic tool across different biofluids and synucleinopathies.

### SAIA Detection of Seeded α-Synuclein in Idiopathic iRBD: Implications for Early Detection

We tested a German cohort of patients with iRBD from the DeNoPa study comprising of 19 iRBD patients and 51 controls. Remarkably, SAIA was able to correctly identify all iRBD samples as test-positive (sensitivity=100%, p<0.001), while significantly discriminating them from controls (specificity=80%, p<0.001) (Figure 6; Supplementary Table 2). The same iRBD samples tested by our SAIA were previously tested in another study using SAA ^54^. Results from SAA study showed 3 of the 19 iRBD cases to be negative and highlighted in Figure 6 as open green triangles (also see Table 1). Two were among the lowest quantified samples, close to the cut-off value (1.7ng/ml), using SAIA. An advantage presented in testing samples from the DeNoPa cohort is the availability of longitudinal clinical information up to 10yrs. This included information on the iRBD cases that had been converted to PD or DLB. The samples tested for this cohort by SAIA were obtained at baseline. Of the 19 iRBD cases tested from this cohort, four patients subsequently converted to DLB (represented as purple colored triangles), while four other iRBD cases converted to PD (represented as orange colored triangles) (Figure 6a, Table 1). Remarkably, the levels of seeded α-synuclein detected in iRBD cases that converted to DLB were lower compared to those that converted to PD. The UK cohort included in this study contained 5 cases of iRBD with CSF samples which we tested. Similar to the German iRBD cohort, all 5 iRBD cases tested positive in SAIA and corresponded as positive by SAA in previously published results (Figure 6b, Table 1) ^26^. Of the 5 iRBD cases, one case later converted to PD and another case converted to pure autonomic failure (PAF). These collective results suggest the reliability of SAIA as a potential early detection method for prodromal α-synucleinopathies.

**Figure 6.**
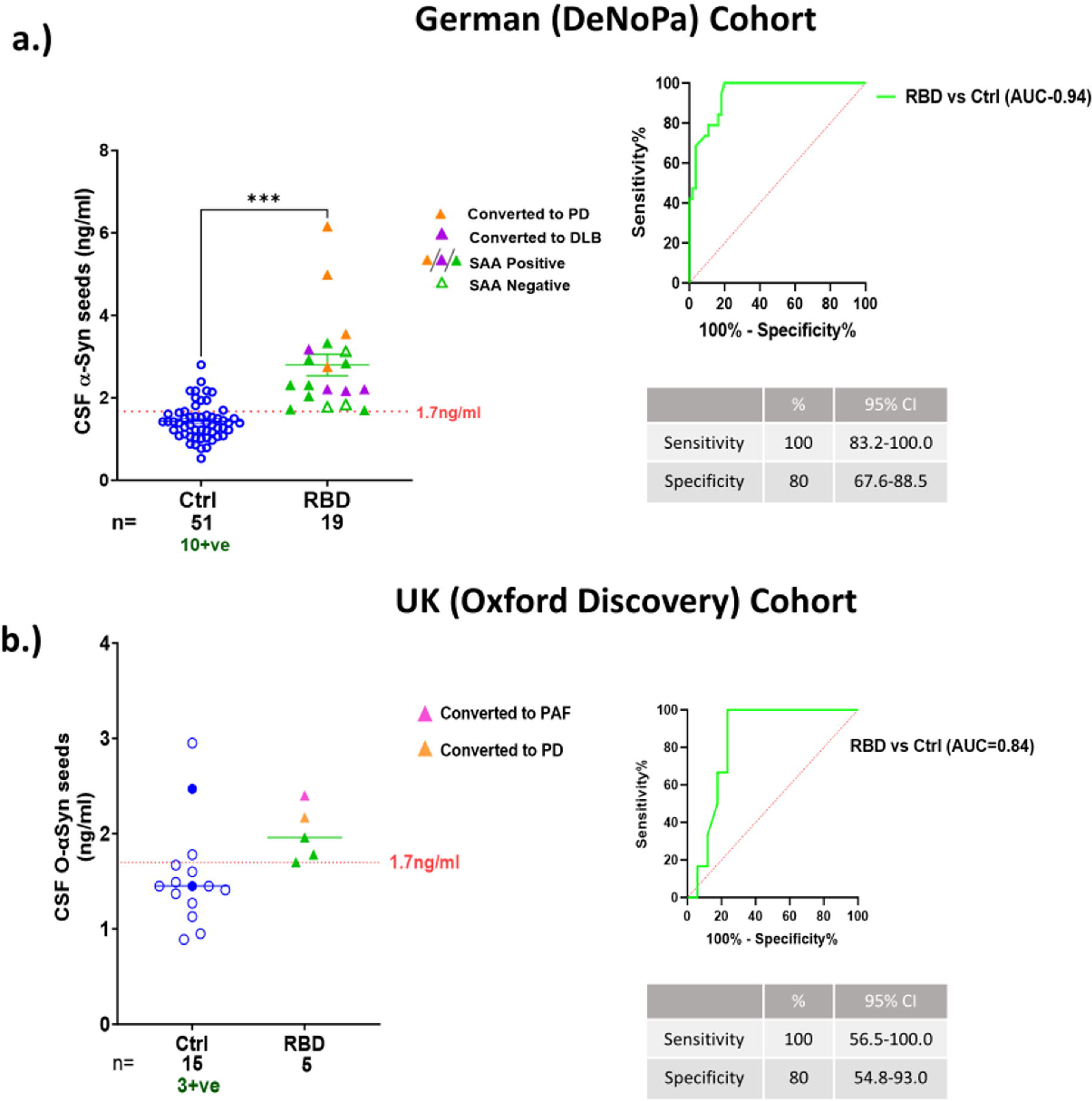
**SAIA’s detection sensitivity in prodromal synucleinopathies cases.** SAIA’s diagnostic performance in detecting prodromal α-synucleinopathies in patients with iRBD is highlighted. Analysis involved two cohorts **a.)** German (DeNoPa) cohort of 19 iRBD patients and 51 healthy controls, and **b.)** UK (Oxford Discovery) cohort of 5 iRBD patients and 15 controls. Scatter plots and ROC curve analyses underline SAIA’s proficiency in quantifying the levels of seeded α-synuclein with a sensitivity of 100% and specificity of 80%. SAIA results and clinical longitudinal data were compared with SAA results and indicated in the graph as: open green triangles/circles- SAA negative samples, orange colored triangles- subjects that converted to PD, purple colored triangles- subjects that converted to DLB, and pink colored triangles-subjects that converted to PAF. *Significant differences between groups were calculated by Man-Whitney test (***- p<0.001, **-p<0.01). Cut-off of 1.7ng/ml was calculated from Youden’s Index from ROC analysis. The error bars represent the standard error of the mean*.

**Table 1.** Comparison of SAA vs SAIA results in iRBD Samples.

| German Cohort (DeNoPa) |  |  |  |  |
| --- | --- | --- | --- | --- |
| Case | SAA results<br>(Positive/Negative) | SAIA results<br>(Conc. ng/ml) | Final Diagnosis | Comment |
| iRBD 1 | Positive | 2.04 | Dementia with Lewy bodies |  |
| iRBD 2 | Positive | 2.21 | Dementia with Lewy bodies |  |
| iRBD 3 | Positive | 3.18 | Dementia with Lewy bodies |  |
| iRBD 4 | Positive | 2.17 | Dementia with Lewy bodies |  |
| iRBD 5 | Positive | 2.21 | No conversion yet |  |
| **iRBD 6** | **Negative** | **1.84** | **No conversion yet** | **drop-out of RBS study: iRBD**<br>**not certain** |
| iRBD 7 | Positive | 3.33 | No conversion yet |  |
| **iRBD 8** | **Negative** | **1.78** | **No conversion yet** | **drop-out of iRBD study: PSG**<br>**interpretation impaired due to**<br>**severe sleep apnea** |
| **iRBD 9** | **Negative** | **3.13** | **No conversion yet** |  |
| iRBD 10 | Positive | 2.93 | No conversion yet |  |
| iRBD 11 | Positive | 2.31 | No conversion yet |  |
| iRBD 12 | Positive | 2.84 | No conversion yet |  |
| iRBD 13 | Positive | 1.80 | No conversion yet |  |
| iRBD 14 | Positive | 2.31 | No conversion yet |  |
| iRBD 15 | Positive | 2.01 | No conversion yet |  |
| iRBD 16 | Positive | 4.99 | Parkinson's Disease |  |
| iRBD 17 | Positive | 6.16 | Parkinson's Disease |  |
| iRBD 18 | Positive | 3.55 | Parkinson's Disease |  |
| iRBD 19 | Positive | 2.75 | Parkinson's Disease |  |
| UK Cohort |  |  |  |  |
| iRBD 1 | Positive | 2.17 | Parkinson's Disease |  |
| iRBD 2 | Positive | 2.40 | Pure autonomic failure |  |
| iRBD 3 | Positive | 1.96 | No conversion yet |  |
| iRBD 4 | Positive | 1.78 | No conversion yet |  |
| iRBD 5 | Positive | 1.70 | No conversion yet |  |

A select number of positive, negative, and all borderline cases from each group were retested to confirm the results obtained. Retest results confirmed original results obtained. We were unable to retest the control samples due to lack of sample availability.

### Evaluating SAIA Specificity in Distinguishing Synucleinopathies from Non-Synucleinopathy Disorders

To assess the specificity of SAIA for synucleinopathies, we analyzed CSF samples from a Greek cohort consisting of PD, PDD, DLB, and MSA patients (n=34, 12, 26, and 36, respectively), as well as non-synucleinopathy including PSP, AD, VaD, and MCI patients (n=34, 46, 16, and 7, respectively- Supplementary Table 1). With the exception of 12 out of 46 (26%) AD cases and 1 VaD case that tested as “positive”, the majority of non-synucleinopathy cases showed a negative result for seeded α-synuclein. Collectively our results demonstrated the high specificity of SAIA for synucleinopathies, with sensitivities ranging from 84% to 100% and specificities of 100% in relation to tauopathy PSP. While significant differences were not observed between the synucleinopathies tested, a trend of lower levels of seeded α-synuclein was observed in DLB cases (mean-2.2±0.6) compared to PD (mean=2.7±0.6), PDD (mean=3.1±1.2), and MSA (mean=2.6±0.6). The AUC values were 0.98 for PD, 1 for PDD, 0.99 for MSA, and 0.91 for DLB, further supporting the discriminative power of SAIA (Figure 7 a-b; Supplementary Table 2).

**Figure 7.**
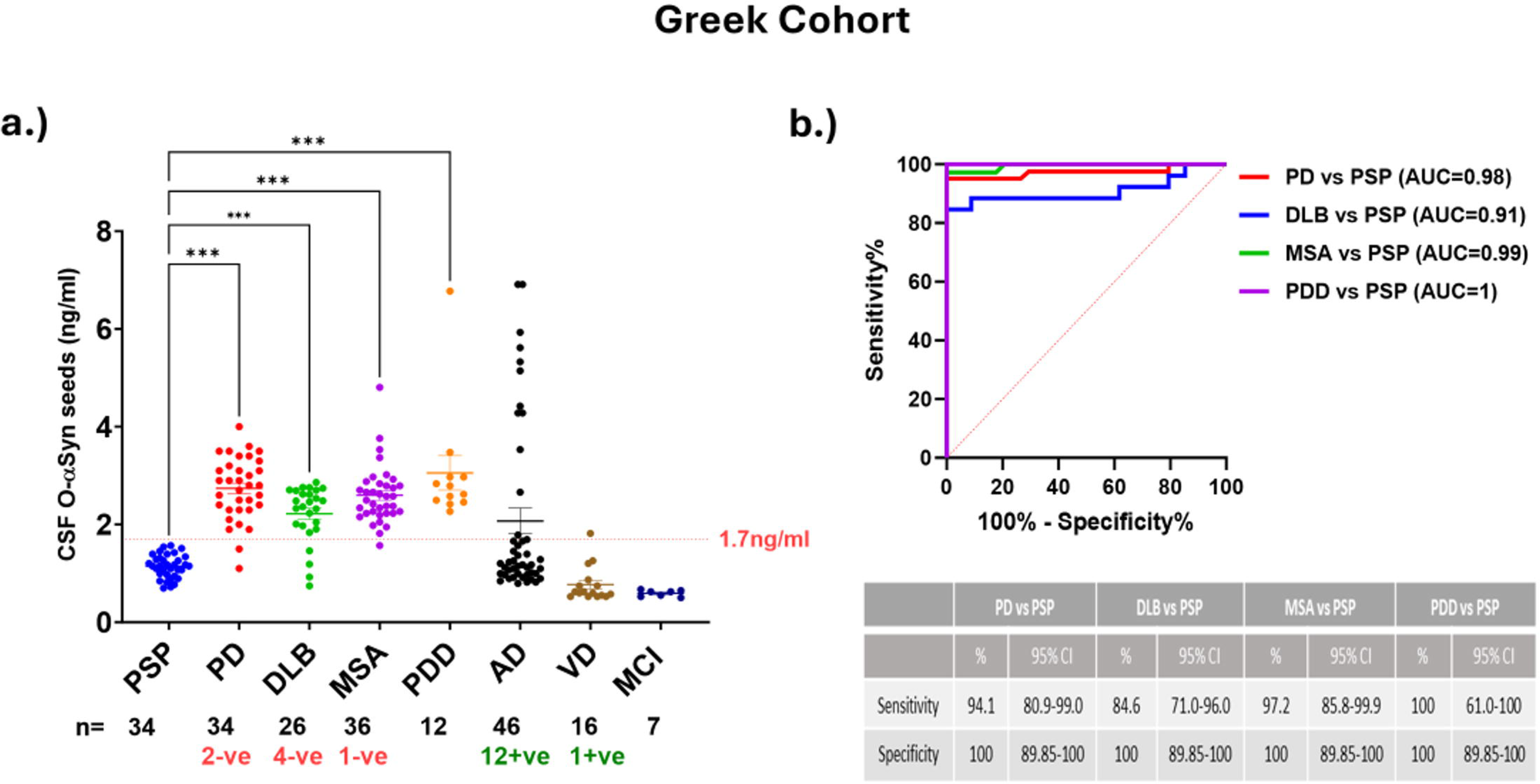
SAIA Specificity in detecting synucleinopathies. **a.**) This figure displays SAIA’s specificity in detecting synucleinopathies. CSF samples from PD, PDD, DLB, and MSA patients (n=41, 6, 26, and 36) are compared with non-synucleinopathy patients with PSP, AD, VaD, or MCI (n=34, 46, 16, and 7). **b.)** ROC curve demonstrating SAIA’s ability to differentiate between PSP, as a control group, and the synucleinopathy groups PD (red line; Sensitivity=94%), DLB (blue line; Sensitivity=85%), MSA (green line; Sensitivity=97%), PDD (purple line; Sensitivity=100%). *Significant differences between groups were calculated by Kruskal- Wallis Test (***- p<0.001, **-p<0.01). Cut-off of 1.7ng/ml was calculated from Youden’s Index from ROC analysis. The error bars represent the standard error of the mean*.

### Improving SAIA Specificity for Distinguishing Between Synucleinopathies

In the initial evaluation of SAIA’s specificity, the assay effectively distinguished synucleinopathies from non-synucleinopathy disorders. However, under standard conditions optimized for SAIA (18 hours incubation), no significant differentiation between specific synucleinopathies, such as PD and MSA, was observed. This was in contrast to previous studies using SAA, which demonstrated the ability to differentiate these groups based on distinct aggregation kinetics. To address this limitation, we optimized SAIA by performing timepoint analyses to determine whether varying incubation times could enhance the assay’s specificity for distinguishing between PD and MSA.

A series of timepoint experiments were conducted using BH samples from MSA and PD patients, with incubations at 2, 4, 6, 8, 14, and 18 hours. MSA cases exhibited rapid aggregation, with detectable seeded α-synuclein as early as 2 hours (MSA mean = 2.02 ± 1.09 vs Ctrl mean = 0.66 ± 0.15, p < 0.05), while PD cases showed a trend towards higher levels (mean = 1.02 ± 0.19, p = 0.12). By the 4 and 6-hour timepoints, significant differences were observed between both MSA and PD compared to controls, with MSA showing a higher trend than PD. By 8 hours, levels began to drop, with a more pronounced reduction at 18 hours, especially in MSA cases (Figure 8a). These findings suggest that prolonged incubation (18 hours) may obscure kinetic differences between MSA and PD, leading to an overlap in α- synuclein seed detection.

**Figure 8.**
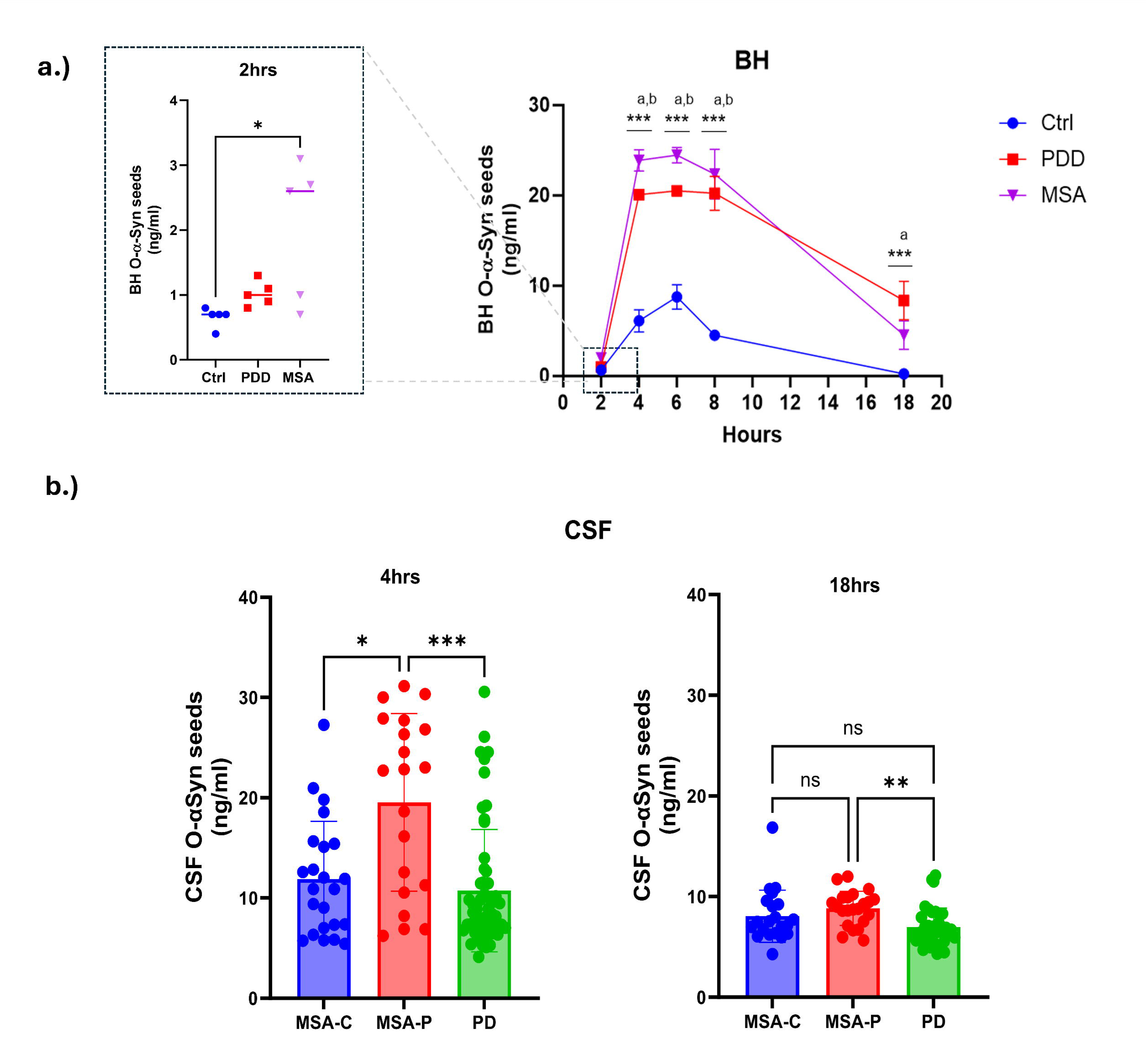
**Timepoint optimization improves SAIA specificity for distinguishing synucleinopathies. *a***.) Aggregation kinetics of α-synuclein seeds in brain homogenate (BH) samples from MSA, PD, and control patients at multiple incubation timepoints (2, 4, 6, 8, and 18 hours). MSA cases (n=5) show significant early aggregation compared to controls (n=5) at 2 hours (p < 0.05), with PD cases (n=5) trending higher but not reaching significance. **b.)** Comparison of α-synuclein seed levels in CSF samples from MSA-C, MSA-P, and PD cases (n=23, 20, and 57, respectively) at 4 hours. MSA-P shows significantly higher levels of seeded α-synuclein compared to MSA-C and PD (p < 0.05 and 0.001 respectively). **c.)** Comparison of α-synuclein seed levels in CSF samples from a smaller Netherlands (Progress- PD) cohort (Ctrl: n=8; PD: n=9) at 4 and 18 hours. At 4 hours, PD cases show significantly higher levels of α-synuclein seeds compared to controls (p < 0.001), a difference that remains robust at 18 hours (p < 0.001). *Significant differences between groups were calculated by Mann-Whitney or Kruskal-Wallis test (*-p<0.01, **- p< 0.05, ***-p<0.001; a-p<0.001 PDD vs Ctrl, b-p<0.001 MSA vs Ctrl). Error bars represent the standard error of the mean*.

To further explore this, we conducted timepoint analyses using CSF samples from a UK cohort (UCL) consisting of MSA (n=43) and PD (n=57) cases. The MSA group included MSA-C (n=23) and MSA-P (n=20) cases. Incubating sample for 4hrs instead of 18hrs showed that MSA-P samples (mean = 19.54 ± 8.9) had significantly higher levels of α- synuclein seeds compared to MSA-C (mean = 11.88 ± 5.76, p < 0.05) and PD cases (mean = 10.74 ± 6.1, p<0.001). suggesting that shorter incubation times enhance SAIA’s specificity for distinguishing MSA from PD.

Given that the earlier cohorts were tested with the longer incubation protocol, we sought to investigate how shorter incubation would affect the results previously achieved for PD. Due to sample availability restriction, we could only test a small number of the Netherlands (Progress-PD) cohort (Ctrl: n=8; PD: n=9). At 4 hours, PD cases showed significantly higher levels of α-synuclein seeds compared to controls (p < 0.001), with a clear separation between the groups. This separation was maintained at 18 hours, where PD cases continued to show significantly elevated α-synuclein levels compared to controls (p < 0.001). Unlike the overlap observed in disease-specific comparisons (e.g., MSA vs PD), these results reinforce the robustness of SAIA for distinguishing PD from controls at both timepoints.

## Discussion

Early and accurate detection of α-synuclein pathology, a hallmark of synucleinopathies, is paramount for effective disease management and intervention. Emerging techniques over recent years, in particular methods such as SAA, have shown considerable promise as a powerful tool to detect misfolded forms of α-synuclein. Moreover, SAA test positivity has entered -as of 2024- a new classification system of PD, which has begun a new era of a biological definition of the disorder for clinical research purposes^55,56^. Several studies utilizing SAA have successfully shown its efficiency in detecting α-synuclein seed activity in BH and CSF samples from patients with PD, DLB, and even MSA reporting high diagnostic sensitivities (exceeding 90%) and specificities ranging of 82–100% ^15,57,58^. Some studies have also adapted the assay for use with other biological samples like nasal brushings, salivary glands, and skin samples^31,59,60^. The assay presents a proficient clinical tool with its high diagnostic potential and its capability for automated monitoring of aggregation kinetics. However, it exhibited limitations in terms of it lacking quantitative capabilities due to its binary nature and robustness due to variations in assay conditions thus hindering its widespread adoption in clinical research and diagnostics. Moreover, its complex technical requirements pose challenges for routine laboratory implementation. The inability to monitor target engagement and α-synuclein aggregation *in vivo* has left a void in the field, as highlighted in recent clinical trials ^61^.

This methodology represents a significant advancement in the field by introducing the creation of a first-in-kind SAIA, which shows a comprehensive investigation of its capabilities harnessing diverse range of cohorts from different populations with synucleinopathies. This approach at once solves the limitations of current diagnostic method SAA by simultaneously amplifying and quantifying α-synuclein seeds within samples and offering a breakthrough in both sensitivity and specificity.

The concept of the assay design was proven in a pilot study of *postmortem* BH and CSF samples. Results of pilot demonstrated the occurrence of α-synuclein seeding specifically in disease cases tested (PD/PDD) samples while control samples remained negative. These results highlight SAIA’s potential diagnostic value and its ability to capture and amplify minimal levels of endogenous α-synuclein seeds in synucleinopathies.

Having established the efficacy of the SAIA method, we conducted a comprehensive evaluation of the assay’s sensitivity and robustness. The assay’s sensitivity was rigorously assessed by testing serial dilutions of BH samples from PDD, DLB and MSA cases and using α-synuclein PFFs in minute quantities. These experiments highlighted the high sensitivity of SAIA, enabling for the accurate detection and quantification of even tiny amounts of α- synuclein seeds. The ability to detect seeded α-synuclein in diluted samples of BH samples in short time of incubation (∼18hrs) from subjects pathologically confirmed with PD, DLB, or MSA up to 6,400 times emphasizes the assay’s potential as a highly sensitive diagnostic tool. Moreover, the successful amplification and quantification of attogram-level quantities of α- synuclein PFFs in control BH and ACSF buffer further underscore the precision and robustness of SAIA, positioning it as an invaluable technique in studying α-synuclein pathology with sensitivity levels comparable to the SAA ^62,63^. This is particularly noted in that the sensitivity achieved with the SAIA is within a short time of only 18hrs of sample incubation using very low concentration of substrate (∼10µg/ml), α-synuclein monomer, as well as similar condition regardless of biofluid tested which is not the case with the SAA which require much higher substrate concentrations (10x-100x more), longer incubation periods (3-7 days), and the use of different protocols for BH and CSF, even requiring beads in some protocols that can create an additional factor for variability. A critical aspect of our study was the assessment of the SAIA assay’s robustness. Robustness in diagnostic assays is defined by their ability to provide consistent, reliable results under varied conditions and across different sample types. In our study, this was evaluated through repeat measurements of both BH and CSF samples, as well as the calibrator standards, on multiple non-consecutive days. The results demonstrated remarkable consistency, with inter- and intra-assay variability observed to be below 15%.

Validation of the SAIA assay in a larger cohort of *postmortem* BH samples, including subjects with PDD, DLB, MSA, and controls, consistently demonstrated higher levels of seeded α-synuclein specifically in synucleinopathy cases. This indicates that SAIA is effective in detecting α-synuclein aggregates in postmortem brain tissue, offering a potential reliable diagnostic tool for α-synucleinopathies. The versatility of SAIA was further demonstrated in its validation with a larger CSF cohort of sporadic PD and control subjects from the UK (Oxford Discovery Cohort). SAIA effectively differentiated PD CSF samples from controls, underscoring its potential as a diagnostic tool for synucleinopathies. Further validation in an additional CSF cohort from the Netherlands reinforced SAIA’s diagnostic potential. Both cohorts showed clear distinctions between PD and control samples, with sensitivities and specificities above 90% and 85%, respectively.

Additionally, we sought to corroborate the findings from the initial pilot study by conducting a side-by-side comparison with the traditional SAA to establish the validity of SAIA in diagnosing cases of synucleinopathies. The SAA and SAIA results in both BH and CSF samples tested largely concurred with each other. As established in the pilot study, the comparison of SAIA results with SAA results in both BH and CSF samples showed a high correlation in the detection rates of synucleinopathy cases. This validation underscores the efficacy of SAIA as a candidate diagnostic assay for synucleinopathies, with the added advantage of quantifying α-synuclein seeds. The inclusion of previously published SAA results^26^ for the UK Oxford Discovery cohort allowed for comparative analysis, revealing some inconsistencies between SAA and SAIA results. Interestingly, SAIA demonstrated potential increased sensitivity compared to SAA, as some CSF PD cases that tested negative with SAA were detectable as positive using SAIA. Specifically, 12 PD cases and 1 control showed discordant results between the two assays. Of the 12 PD cases, 8 were SAIA-positive but SAA-negative, while the other 4 showed the opposite pattern. These discrepancies were confirmed by repeated testing with both assays to confirm the results through repeated testing with SAIA, suggesting that while SAIA generally aligns with SAA results, it may offer additional diagnostic insights or detect α-synuclein aggregates that the Th-T dye in SAA might miss. The observed correlations between SAIA-seeded α-synuclein levels and SAA kinetic parameters suggest that SAIA may provide results comparable to SAA in reflecting α- synuclein aggregation dynamics. The presence of discordant samples underscores the need for further investigation into factors that may influence assay outcomes, such as differences in aggregation properties or sample composition.

Expanding the scope of SAIA, the assay’s performance was evaluated in prodromal cases of PD/DLB, specifically iRBD. SAIA successfully detected seeded α-synuclein in all iRBD CSF samples, offering promise for early-stage synucleinopathy diagnosis. Notably, our study’s results align with those previously published using SAA^16,25,64,65^. Moreover, the availability of longitudinal data for the German (DeNoPa) and UK (Oxford Discovery) cohorts as well as SAA data for the same samples tested adds significant value to our findings. The SAA results showed three negative iRBD samples that tested positive with SAIA, two of which were borderline positive. Longitudinal clinical cohort information showed that four out of the 19 iRBD patients from the German cohort progressed to DLB, and another four developed PD. Two iRBD patients from the UK cohort had also converted to PD and PAF, a rarer type of α-synucleinopathy. These transitions, indicated in Table 1, underline the potential of SAIA not just as a diagnostic tool but also as a predictor for the progression of iRBD to definitive synucleinopathies. Also noted 3 out of the four iRBD subjects that converted to PD measured the highest levels of seeded α-synuclein compared to non-converted iRBD and to those converted to DLB. This could potentially be noteworthy observation given the trend of lower levels of CSF seeded α-synuclein measured in DLB cases compared to PD in our study in the Greek cohort. The consistent detection of iRBD cases across different cohorts reinforces the robustness of SAIA as an early diagnostic tool for prodromal α-synucleinopathies. However, we acknowledge that further validation through larger longitudinal studies would be invaluable in fully establishing its clinical utility in this context.

Importantly, SAIA demonstrated specificity for synucleinopathies when analyzing CSF samples from various neurodegenerative diseases, with the majority of non-synucleinopathy cases yielding negative results except for 12 out of 46 (26%) AD cases that tested positive. These findings align with previous reports of α-synuclein pathology in a subset of AD patients at autopsy ^40,66–68^, and are consistent with prior research conducted using the conventional SAA method ^8,16,25,62,69^. It is worth noting that the absence of α-synuclein pathology in cases of MCI tested, often considered as prodromal cases of AD, suggests that α-synuclein pathology may emerge later in the course of dementia as a secondary pathology. This aligns with studies, such as Levin *et al*.^70^, that have shown α-synuclein aggregates are typically detected late in the disease course and depend on the Lewy pathology burden. The lower detection rate in our AD cohort may reflect the early stage of disease in some patients, where the α-synuclein burden remains low and more difficult to detect.

Interestingly, the α-synuclein levels detected in some positive AD cases were higher than those observed in PD, PDD, or DLB patients. This observation may reflect variability in the α-synuclein species present in AD patients. Post-mortem studies have shown that AD cases can exhibit distinct molecular signatures of α-synuclein compared to those found in synucleinopathies like PD or DLB^71^. These species-specific differences may influence the SAIA assay’s ability to detect and quantify α-synuclein aggregates, potentially leading to the higher levels observed in some AD patients. While the exact reasons for these elevated levels remain unclear, this variability underscores the complexity of α-synuclein pathology across different neurodegenerative diseases.

Additionally, the assay’s design focuses on measuring soluble α-synuclein seeds, which could further explain the discrepancies between our results and studies that assess insoluble aggregates, such as Lewy bodies. In AD, α-synuclein pathology may still be in an earlier phase, with more soluble species present, while in DLB and PD, more α-synuclein could have progressed to insoluble aggregates, which are less detectable by SAIA. This might account for the lower seed concentrations observed in DLB compared to AD, despite the higher Lewy body burden typically seen in DLB. The differences in seed concentrations observed across diseases highlight the complexity of α-synuclein pathology and suggest that SAIA’s focus on soluble seeds may offer a complementary perspective to studies focusing on insoluble aggregates. While these explanations remain speculative, they offer a framework for understanding the unexpected results, and further research will be needed to explore the relationship between soluble and insoluble α-synuclein species and disease progression. No α-synuclein pathology was detected in the PSP cohort, despite previous reports indicating a presence of α-synuclein pathology in approximately 10-20% in post-mortem studies ^72,73^. Diagnosing PSP in living patients is inherently challenging due to clinical overlaps with other neurodegenerative disorders and the variability of its presentations. The patients included in our study met the 2017 Movement Disorder Society (MDS) diagnostic criteria for PSP. While these criteria improve specificity, the sensitivity for detecting variant forms of PSP remains limited, which could explain the discrepancies between our findings and post-mortem data.

SAIA could not significantly differentiate between these distinct synucleinopathies tested in this study, however a trend of lower levels of seeded α-synuclein in DLB cases compared to PD, PDD, and MSA was observed. We could also speculate that the lack of differentiation could be due to the strain of α-synuclein captured by the conformation-specific Ab, 2A1 which detects C-terminally truncated as well as full-length α-synuclein seeds. Several studies have demonstrated that there are differences in the strains of α-synuclein present in different α-synucleinopathies with some cases also indicating heterogeneity of species found ^57,74–77^. Nonetheless SAIA technique holds potential for enhanced sensitivity to better stratify and distinguish between these diseases through the use of other strain-specific anti-α-synuclein antibodies, different reaction buffer ^24,57^, truncated α-synuclein as substrate ^78^ or even possibility exploring combining SAIA results with other methods, such as autonomic function testing, neuroimaging, and/or genetics ^79–81^.

One limitation observed in this study was the lack of clear differentiation between PD and MSA under the standard 18-hour incubation time, despite SAA’s ability to distinguish these diseases based on aggregation kinetics. To address this, we explored the effects of shorter incubation times on SAIA’s performance. Timepoint experiments revealed that BHs from MSA cases exhibited significantly faster aggregation, with detectable α-synuclein seeds as early as 2 hours, while PD cases showed slower aggregation, reaching significant levels at 6 to 8 hours. By the 18-hour timepoint, the signal in both groups had declined, particularly in MSA cases, resulting in an overlap between PD and MSA. This reduction in signal may be attributed to the assay’s design, which targets soluble α-synuclein seeds, with the later aggregation phases potentially forming insoluble aggregates that are less detectable by SAIA.

In CSF samples, a similar pattern emerged. At 4 hours, a clear distinction between MSA-P and PD was observed, with MSA-P showing significantly higher levels of α-synuclein seeds compared to both MSA-C and PD. This difference in results observed between MSA-P and MSA-C aligns with findings from Bargar *et al*.^82^, who demonstrated a lack of seeding activity in olfactory mucosa samples from MSA-C patients, in contrast to the pronounced seeding activity observed in MSA-P cases using SAA. Their study found that MSA-C cases were mostly negative, whereas MSA-P cases were largely positive, a finding consistent with the more robust aggregation we observed in MSA-P at earlier timepoints. Although our primary aim was to differentiate between these MSA and PD groups, the incidentally observed significant differences between MSA-C and MSA-P suggest that the underlying pathology of the two sub-groups may involve different α-synuclein species or aggregation kinetics, which may not be captured at later incubation times or by focusing solely on insoluble aggregates. However, further studies are necessary to validate these findings.

To determine how shorter incubation times might influence our earlier findings, where samples were incubated for 18 hours, we reanalyzed a small number of CSF samples from the Netherlands cohort at 4 hours. PD cases showed significantly elevated α-synuclein seed levels compared to controls, and this difference remained robust at 18 hours. These results demonstrate that both shorter (4 hours) and longer (18 hours) incubation times are effective for distinguishing disease cases (PD) from controls, with no evidence of overlap between groups. Furthermore, our findings highlight that shorter incubation times may enhance SAIA’s specificity for distinguishing between synucleinopathies, such as MSA and PD, particularly during the early aggregation phases when soluble α-synuclein seeds dominate. This optimized approach offers a foundation for refining SAIA’s diagnostic applications, improving its clinical utility for disease-specific diagnosis and providing valuable tools for emerging pharmacological treatments and clinical trials targeting specific MSA subtypes. In summary, these results emphasize the flexibility of SAIA in addressing different diagnostic questions. While longer incubation times may maximize sensitivity for distinguishing disease cases from controls, shorter incubation times appear more effective for capturing disease-specific differences between synucleinopathies. Further studies across additional cohorts and incubation parameters will be critical to validate and expand these findings.

We acknowledge that this study has limitations. In the iRBD cohort tested, the specificity rate (∼80%) obtained was lower than other cohorts tested due to a positive test result in select controls. We could speculate however that these positive controls possibly harbored incidental Lewy body disease. Given the sensitivity of the assay to low levels of α-synuclein seeds, it is possible that SAIA could detect and amplify the minimal amount of α-synuclein seeds present in incidental Lewy bodies which have been indicated be found in 14% of the aging population (range 40-100 years) ^83^. Also needed are longitudinal studies to assess clinical correlations, as well as evaluating target engagement in clinical trials.

In summary, we have developed a quantitative high throughput immunoassay for synucleinopathies with high diagnostic potential for use in diagnostics, longitudinal studies, and clinical trials. SAIA represents a significant methodological advance, simultaneously amplifying and quantifying α-synuclein seeds, offering a streamlined approach for diagnosing, monitoring, and assessing drug engagement in synucleinopathies. The assay’s high sensitivity, specificity, and versatility make it a promising tool for the detection, quantification, and monitoring α-synuclein pathology *in vivo*. Importantly, this simplified assay can be readily adopted in most clinical and research laboratories, as it relies on a traditional colorimetric ELISA platform. This eliminates the need for sophisticated and costly equipment or specialized laboratories requiring extensive technical expertise.

## Data availability

The datasets used and/or analyzed during the current study are available from the corresponding author on reasonable request.

## Acknowledgements

The authors express their sincere gratitude to all the participants who generously contributed their samples for this study. They also extend our appreciation to the funding agencies and institutions that supported this research, enabling the advancement of our understanding of synucleinopathies and the development of SAIA.

---

Supplementary Figure 1. **Epitope mapping and isotyping of 2A1 antibody** (a) Epitope mapping analysis using α-synuclein peptides, demonstrating that 2A1 specifically binds to peptide 17, corresponding to amino acids 113–123 of α-synuclein. (b) Isotyping analysis confirming that 2A1 is an IgG2a subclass antibody with a kappa light chain.

Supplementary Figure 2. **Correlation between** α**-synuclein seeding activity and Lewy body burden in brain homogenates from the Newcastle cohort.** Scatter plots show the relationship between α-synuclein seeding activity measured by SAIA and the mean percentage area of Lewy body (LB) staining in **(a)** all cases combined (PDD and DLB), **(b)** PDD cases alone, and **(c)** DLB cases alone. A significant positive correlation is observed in PDD cases (b, red squares, r = 0.75, p < 0.05), while no correlation is detected in DLB cases (c, green triangles, r = -0.30, p = 0.40). Linear regression lines are shown for each group.

## References

1. Barker RA, Williams-Gray CH. Review: The spectrum of clinical features seen with alpha synuclein pathology. Neuropathol Appl Neurobiol. 2016;42(1):6–19. doi:10.1111/NAN.12303

2. Goedert M, Jakes R, Spillantini MG. The Synucleinopathies: Twenty Years On. J Parkinsons Dis. 2017;7(Suppl 1):S51. doi:10.3233/JPD-179005

3. Fayyad M, Salim S, Majbour N, et al. Parkinson’s disease biomarkers based on α- synuclein. J Neurochem. 2019;150(5):626–636. doi:10.1111/JNC.14809

4. Youssef P, Kim WS, Halliday GM, Lewis SJG, Dzamko N. Comparison of Different Platform Immunoassays for the Measurement of Plasma Alpha-Synuclein in Parkinson’s Disease Patients. J Parkinsons Dis. 2021;11(4):1761. doi:10.3233/JPD-212694

5. Tsao HH, Huang CG, Wu YR. Detection and assessment of alpha-synuclein in Parkinson disease. Neurochem Int. 2022;158:105358. doi:10.1016/J.NEUINT.2022.105358

6. Majbour NK, Vaikath NN, van Dijk KD, et al. Oligomeric and phosphorylated alpha-synuclein as potential CSF biomarkers for Parkinson’s disease. Mol Neurodegener. 2016;11(1):7. doi:10.1186/s13024-016-0072-9

7. Anagnostou D, Sfakianaki G, Melachroinou K, et al. Assessment of Aggregated and Exosome-Associated α-Synuclein in Brain Tissue and Cerebrospinal Fluid Using Specific Immunoassays. Diagnostics 2023, Vol 13, *Page* 2192. 2023;13(13):2192. doi:10.3390/DIAGNOSTICS13132192

8. Fairfoul G, McGuire LI, Pal S, et al. Alpha-synuclein RT-QuIC in the CSF of patients with alpha-synucleinopathies. Ann Clin Transl Neurol. 2016;3(10):812–818. doi:10.1002/ACN3.338

9. Shahnawaz M, Tokuda T, Waraga M, et al. Development of a Biochemical Diagnosis of Parkinson Disease by Detection of α-Synuclein Misfolded Aggregates in Cerebrospinal Fluid. JAMA Neurol. 2017;74(2):163–172. doi:10.1001/JAMANEUROL.2016.4547

10. Fayyad M, Majbour NK, Vaikath NN, et al. Generation of monoclonal antibodies against phosphorylated α-Synuclein at serine 129: Research tools for synucleinopathies. Neurosci Lett. 2020;725:134899. 10.1016/j.neulet.2020.134899

11. Majbour NK, Chiasserini D, Vaikath NN, et al. Increased levels of CSF total but not oligomeric or phosphorylated forms of alpha-synuclein in patients diagnosed with probable Alzheimer’s disease. Sci Rep. 2017;7. doi:10.1038/srep40263

12. Majbour NK, Vaikath NN, Eusebi P, et al. Longitudinal changes in CSF alpha-synuclein species reflect Parkinson’s disease progression. Movement Disorders. 2016;31(10):1535–1542. doi:10.1002/mds.26754

13. Majbour NK, Abdi IY, Dakna M, et al. Cerebrospinal α-Synuclein Oligomers Reflect Disease Motor Severity in DeNoPa Longitudinal Cohort. Movement Disorders. 2021;36(9):2048–2056. doi:10.1002/MDS.28611

14. Majbour NK, Aasly JO, Hustad E, et al. CSF total and oligomeric α-Synuclein along with TNF-α as risk biomarkers for Parkinson’s disease: a study in LRRK2 mutation carriers. Transl Neurodegener. 2020;9(1):15. doi:10.1186/s40035-020-00192-4

15. Grossauer A, Hemicker G, Krismer F, et al. α Synuclein Seed Amplification Assays in the Diagnosis of Synucleinopathies Using Cerebrospinal Fluid—A Systematic Review and Meta Analysis. Mov Disord Clin Pract. 2023;10(5):737. doi:10.1002/MDC3.13710

16. Rossi M, Candelise N, Baiardi S, et al. Ultrasensitive RT-QuIC assay with high sensitivity and specificity for Lewy body-associated synucleinopathies. Acta Neuropathol. 2020;140(1):49–62. doi:10.1007/S00401-020-02160-8/FIGURES/3

17. Nakagaki T, Nishida N, Satoh K. Development of α-Synuclein Real-Time Quaking-Induced Conversion as a Diagnostic Method for α-Synucleinopathies. Front Aging Neurosci. 2021;13:703984. doi:10.3389/FNAGI.2021.703984/BIBTEX

18. Russo MJ, Orru CD, Concha-Marambio L, et al. High diagnostic performance of independent alpha-synuclein seed amplification assays for detection of early Parkinson’s disease. Acta Neuropathol Commun. 2021;9(1):1–13. doi:10.1186/S40478-021-01282-8/FIGURES/3

19. Srivastava A, Alam P, Caughey B. RT-QuIC and Related Assays for Detecting and Quantifying Prion-like Pathological Seeds of α-Synuclein. Biomolecules. 2022;12(4). doi:10.3390/BIOM12040576

20. Okuzumi A, Hatano T, Matsumoto G, et al. Propagative α-synuclein seeds as serum biomarkers for synucleinopathies. Nature Medicine 2023. Published online May 29, 2023:1-8. doi:10.1038/s41591-023-02358-9

21. Barria MA, Gonzalez-Romero D, Soto C. Cyclic Amplification of Prion Protein Misfolding. Methods Mol Biol. 2012;849:199. doi:10.1007/978-1-61779-551-0_14

22. Concha-Marambio L, Shahnawaz M, Soto C. Detection of Misfolded α-Synuclein Aggregates in Cerebrospinal Fluid by the Protein Misfolding Cyclic Amplification Platform. Methods Mol Biol. 2019;1948:35–44. doi:10.1007/978-1-4939-9124-2_4

23. Saborio GP, Permanne B, Soto C. Sensitive detection of pathological prion protein by cyclic amplification of protein misfolding. Nature. 2001;411(6839):810-813. doi:10.1038/35081095

24. Bellomo G, Paciotti S, Gatticchi L, et al. Seed amplification assays for diagnosing synucleinopathies: The issue of influencing factors. Frontiers in Bioscience - Landmark. 2021;26(11):1075–1088. doi:10.52586/5010/5010/FIG8.JPG

25. Siderowf A, Concha-Marambio L, Lafontant DE, et al. Assessment of heterogeneity among participants in the Parkinson’s Progression Markers Initiative cohort using α- synuclein seed amplification: a cross-sectional study. Lancet Neurol. 2023;22(5):407–417. doi:10.1016/S1474-4422(23)00109-6

26. Poggiolini I, Gupta V, Lawton M, et al. Diagnostic value of cerebrospinal fluid alpha-synuclein seed quantification in synucleinopathies. Brain. 2022;145(2):584–595. doi:10.1093/BRAIN/AWAB431

27. Manne S, Kondru N, Jin H, et al. Blinded RT-QuIC Analysis of α-Synuclein Biomarker in Skin Tissue From Parkinson’s Disease Patients. Movement Disorders. 2020;35(12):2230–2239. doi:10.1002/MDS.28242

28. Manne S, Kondru N, Hepker M, et al. Ultrasensitive Detection of Aggregated α- Synuclein in Glial Cells, Human Cerebrospinal Fluid, and Brain Tissue Using the RT-QuIC Assay: New High-Throughput Neuroimmune Biomarker Assay for Parkinsonian Disorders. J Neuroimmune Pharmacol. 2019;14(3):423–435. doi:10.1007/S11481-019-09835-4

29. Manne S, Kondru N, Jin H, et al. α-Synuclein real-time quaking-induced conversion in the submandibular glands of Parkinson’s disease patients. Mov Disord. 2020;35(2):268–278. doi:10.1002/MDS.27907

30. Bargar C, Wang W, Gunzler SA, et al. Streamlined alpha-synuclein RT-QuIC assay for various biospecimens in Parkinson’s disease and dementia with Lewy bodies. Acta Neuropathol Commun. 2021;9(1):1–13. doi:10.1186/S40478-021-01175-W/FIGURES/5

31. De Luca CMG, Elia AE, Portaleone SM, et al. Efficient RT-QuIC seeding activity for α-synuclein in olfactory mucosa samples of patients with Parkinson’s disease and multiple system atrophy. Transl Neurodegener. 2019;8(1):1–14. doi:10.1186/S40035-019-0164-X/FIGURES/4

32. Bräuer S, Rossi M, Sajapin J, et al. Kinetic parameters of alpha-synuclein seed amplification assay correlate with cognitive impairment in patients with Lewy body disorders. Acta Neuropathol Commun. 2023;11(1):1–9. doi:10.1186/S40478-023-01653-3/FIGURES/3

33. Rossi M, Baiardi S, Teunissen CE, et al. Diagnostic Value of the CSF α-Synuclein Real-Time Quaking-Induced Conversion Assay at the Prodromal MCI Stage of Dementia With Lewy Bodies. Neurology. 2021;97(9):E930–E940. doi:10.1212/WNL.0000000000012438

34. Jensen PH, Schlossmacher MG, Stefanis L. Who Ever Said It Would Be Easy? Reflecting on Two Clinical Trials Targeting α-Synuclein. Movement Disorders. 2023;38(3):378–384. doi:10.1002/MDS.29318

35. Petricca L, Chiki N, Hanna-El-Daher L, et al. Comparative analysis of total alpha-synuclein (αSYN) immunoassays reveals that they do not capture the diversity of modified αSYN proteoforms. bioRxiv. Published online March 14, 2022:2022.03.11.483980. doi:10.1101/2022.03.11.483980

36. Cariulo C, Martufi P, Verani M, et al. Phospho-S129 Alpha-Synuclein Is Present in Human Plasma but Not in Cerebrospinal Fluid as Determined by an Ultrasensitive Immunoassay. Front Neurosci. 2019;13:482808. doi:10.3389/FNINS.2019.00889/BIBTEX

37. Bellomo G, Paciotti S, Gatticchi L, et al. Seed amplification assays for diagnosing synucleinopathies: The issue of influencing factors. Frontiers in Bioscience - Landmark. 2021;26(11):1075–1088. doi:10.52586/5010/5010/FIG8.JPG

38. Majbour N, Aasly J, Abdi I, et al. Disease-Associated α-Synuclein Aggregates as Biomarkers of Parkinson Disease Clinical Stage. Neurology. 2022;99(21):E2417–E2427. doi:10.1212/WNL.0000000000201199

39. Van Dijk KD, Jongbloed W, Heijst JA, et al. Cerebrospinal fluid and plasma clusterin levels in Parkinson’s disease. Parkinsonism Relat Disord. 2013;19(12):1079–1083. doi:10.1016/J.PARKRELDIS.2013.07.016

40. Quadalti C, Palmqvist S, Hall S, et al. Clinical effects of Lewy body pathology in cognitively impaired individuals. Nature Medicine 2023. Published online July 18, 2023:1-7. doi:10.1038/s41591-023-02449-7

41. Hughes AJ, Daniel SE, Kilford L, Lees AJ. Accuracy of clinical diagnosis of idiopathic Parkinson’s disease: a clinico-pathological study of 100 cases. J Neurol Neurosurg Psychiatry. 1992;55(3):181–184. doi:10.1136/JNNP.55.3.181

42. McKeith IG, Boeve BF, DIckson DW, et al. Diagnosis and management of dementia with Lewy bodies: Fourth consensus report of the DLB Consortium. Neurology. 2017;89(1):88–100. doi:10.1212/WNL.0000000000004058

43. Emre M, Aarsland D, Brown R, et al. Clinical diagnostic criteria for dementia associated with Parkinson’s disease. Mov Disord. 2007;22(12):1689–1707. doi:10.1002/MDS.21507

44. Gilman S, Wenning GK, Low PA, et al. Second consensus statement on the diagnosis of multiple system atrophy. Neurology. 2008;71(9):670–676. doi:10.1212/01.WNL.0000324625.00404.15

45. McKhann GM, Knopman DS, Chertkow H, et al. The diagnosis of dementia due to Alzheimer’s disease: recommendations from the National Institute on Aging-Alzheimer’s Association workgroups on diagnostic guidelines for Alzheimer’s disease. Alzheimers Dement. 2011;7(3):263–269. doi:10.1016/J.JALZ.2011.03.005

46. Höglinger GU, Respondek G, Stamelou M, et al. Clinical diagnosis of progressive supranuclear palsy: The movement disorder society criteria. Mov Disord. 2017;32(6):853–864. doi:10.1002/MDS.26987

47. Albert MS, DeKosky ST, Dickson D, et al. The diagnosis of mild cognitive impairment due to Alzheimer’s disease: recommendations from the National Institute on Aging-Alzheimer’s Association workgroups on diagnostic guidelines for Alzheimer’s disease. Alzheimers Dement. 2011;7(3):270–279. doi:10.1016/J.JALZ.2011.03.008

48. Sachdev P, Kalaria R, O’Brien J, et al. Diagnostic criteria for vascular cognitive disorders: a VASCOG statement. Alzheimer Dis Assoc Disord. 2014;28(3):206–218. doi:10.1097/WAD.0000000000000034

49. Postuma RB, Berg D, Stern M, et al. MDS clinical diagnostic criteria for Parkinson’s disease. Movement Disorders. 2015;30(12):1591–1601. doi:10.1002/MDS.26424

50. Szewczyk-Krolikowski K, Tomlinson P, Nithi K, et al. The influence of age and gender on motor and non-motor features of early Parkinson’s disease: Initial findings from the Oxford Parkinson Disease Center (OPDC) discovery cohort. Parkinsonism Relat Disord. 2014;20(1):99–105. doi:10.1016/J.PARKRELDIS.2013.09.025

51. Street D, Jabbari E, Costantini A, et al. Progression of atypical parkinsonian syndromes: PROSPECT-M-UK study implications for clinical trials. Brain. 2023;146(8):3232–3242. doi:10.1093/BRAIN/AWAD105

52. Athauda D, Maclagan K, Skene SS, et al. Exenatide once weekly versus placebo in Parkinson’s disease: a randomised, double-blind, placebo-controlled trial. Lancet. 2017;390(10103):1664. doi:10.1016/S0140-6736(17)31585-4

53. Vaikath N, Sudhakaran I, Abdi I, et al. Structural and Biophysical Characterization of Stable Alpha-Synuclein Oligomers. Int J Mol Sci. 2022;23(23). doi:10.3390/IJMS232314630

54. Concha-Marambio L, Weber S, Farris CM, et al. Accurate detection of α-Synuclein seeds in CSF from iRBD and Parkinson patients in the DeNoPa cohort. Mov Disord. 2023;38(4):567. doi:10.1002/MDS.29329

55. Höglinger GU, Adler CH, Berg D, et al. A biological classification of Parkinson’s disease: the SynNeurGe research diagnostic criteria. Lancet Neurol. 2024;23(2):191–204. doi:10.1016/S1474-4422(23)00404-0

56. Simuni T, Chahine LM, Poston K, et al. A biological definition of neuronal α- synuclein disease: towards an integrated staging system for research. Lancet Neurol. 2024;23(2):178–190. doi:10.1016/S1474-4422(23)00405-2

57. Martinez-Valbuena I, Visanji NP, Kim A, et al. Alpha-synuclein seeding shows a wide heterogeneity in multiple system atrophy. Transl Neurodegener. 2022;11(1). doi:10.1186/S40035-022-00283-4

58. Srivastava A, Alam P, Caughey B. RT-QuIC and Related Assays for Detecting and Quantifying Prion-like Pathological Seeds of α-Synuclein. Biomolecules. 2022;12(4). doi:10.3390/BIOM12040576

59. Kuzkina A, Bargar C, Schmitt D, et al. Diagnostic value of skin RT-QuIC in Parkinson’s disease: a two-laboratory study. npj Parkinson’s Disease 2021 7:*1*. 2021;7(1):1-11. doi:10.1038/s41531-021-00242-2

60. Luan M, Sun Y, Chen J, et al. Diagnostic Value of Salivary Real-Time Quaking-Induced Conversion in Parkinson’s Disease and Multiple System Atrophy. Mov Disord. 2022;37(5):1059–1063. doi:10.1002/MDS.28976

61. Jensen PH, Schlossmacher MG, Stefanis L. Who Ever Said It Would Be Easy? Reflecting on Two Clinical Trials Targeting α-Synuclein. Mov Disord. 2023;38(3):378–384. doi:10.1002/MDS.29318

62. Groveman BR, Orrù CD, Hughson AG, et al. Rapid and ultra-sensitive quantitation of disease-associated α-synuclein seeds in brain and cerebrospinal fluid by αSyn RT-QuIC. Acta Neuropathol Commun. 2018;6. doi:10.1186/S40478-018-0508-2

63. Russo MJ, Orru CD, Concha-Marambio L, et al. High diagnostic performance of independent alpha-synuclein seed amplification assays for detection of early Parkinson’s disease. Acta Neuropathol Commun. 2021;9(1):1–13. doi:10.1186/S40478-021-01282-8/FIGURES/3

64. Iranzo A, Fairfoul G, Ayudhaya ACN, et al. Detection of α-synuclein in CSF by RT-QuIC in patients with isolated rapid-eye-movement sleep behaviour disorder: a longitudinal observational study. Lancet Neurol. 2021;20(3):203–212. doi:10.1016/S1474-4422(20)30449-X

65. Poggiolini I, Gupta V, Lawton M, et al. Diagnostic value of cerebrospinal fluid alpha-synuclein seed quantification in synucleinopathies. Brain. 2022;145(2):584–595. doi:10.1093/BRAIN/AWAB431

66. Berge G, Sando SB, Albrektsen G, et al. Alpha-synuclein measured in cerebrospinal fluid from patients with Alzheimer’s disease, mild cognitive impairment, or healthy controls: a two year follow-up study. BMC Neurol. 2016;16(1). doi:10.1186/S12883-016-0706-0

67. Twohig D, Nielsen HM. α-synuclein in the pathophysiology of Alzheimer’s disease. Molecular Neurodegeneration 2019 14:*1*. 2019;14(1):1-19. doi:10.1186/S13024-019-0320-X

68. Palmqvist S, Rossi M, Hall S, et al. Cognitive effects of Lewy body pathology in clinically unimpaired individuals. Nature Medicine 2023. Published online July 18, 2023:1-8. doi:10.1038/s41591-023-02450-0

69. Concha-Marambio L, Shahnawaz M, Soto C. Detection of misfolded α-synuclein aggregates in cerebrospinal fluid by the protein misfolding cyclic amplification platform. Methods in Molecular Biology. 2019;1948:35–44. doi:10.1007/978-1-4939-9124-2_4/COVER

70. Levin J, Baiardi S, Quadalti C, et al. α-Synuclein seed amplification assay detects Lewy body co-pathology in autosomal dominant Alzheimer’s disease late in the disease course and dependent on Lewy pathology burden. Alzheimers Dement. 2024;20(6):4351–4365. doi:10.1002/ALZ.13818

71. van der Gaag BL, Deshayes NAC, Breve JJP, et al. Distinct tau and alpha-synuclein molecular signatures in Alzheimer’s disease with and without Lewy bodies and Parkinson’s disease with dementia. Acta Neuropathol. 2024;147(1):1–22. doi:10.1007/S00401-023-02657-Y/FIGURES/10

72. Mori H, Oda M, Komori T, et al. Lewy bodies in progressive supranuclear palsy. Acta Neuropathol. 2002;104(3):273–278. doi:10.1007/S00401-002-0555-3/METRICS

73. Aguirre MEE, Zelaya MV, de Gordoa JSR, Tuñón MT, Lanciego JL. Midbrain catecholaminergic neurons co-express α-synuclein and tau in progressive supranuclear palsy. Front Neuroanat. 2015;9(MAR):132680. doi:10.3389/FNANA.2015.00025/BIBTEX

74. Schweighauser M, Shi Y, Tarutani A, et al. Structures of α-Synuclein Filaments from Multiple System Atrophy. Nature. 2020;585(7825):464. doi:10.1038/S41586-020-2317-6

75. Shahnawaz M, Mukherjee A, Pritzkow S, et al. Discriminating α-synuclein strains in Parkinson’s disease and multiple system atrophy. Nature. 2020;578(7794):273-277. doi:10.1038/S41586-020-1984-7

76. Zhou Z, Kihara AH, Graves NJ, Gambin Y, Sierecki E. α-Synuclein Strains and Their Relevance to Parkinson’s Disease, Multiple System Atrophy, and Dementia with Lewy Bodies. International Journal of Molecular Sciences 2023, Vol 24, *Page 12134*. 2023;24(15):12134. doi:10.3390/IJMS241512134

77. Van der Perren A, Gelders G, Fenyi A, et al. The structural differences between patient-derived α-synuclein strains dictate characteristics of Parkinson’s disease, multiple system atrophy and dementia with Lewy bodies. Acta Neuropathol. 2020;139(6):977. doi:10.1007/S00401-020-02157-3

78. Poggiolini I, Erskine D, Vaikath NN, et al. RT-QuIC Using C-Terminally Truncated α- Synuclein Forms Detects Differences in Seeding Propensity of Different Brain Regions from Synucleinopathies. Biomolecules. 2021;11(6). doi:10.3390/BIOM11060820

79. Chahine LM, Merchant K, Siderowf A, et al. Proposal for a Biologic Staging System of Parkinson’s Disease. J Parkinsons Dis. 2023;13(3):297–309. doi:10.3233/JPD-225111

80. Bellomo G, De Luca CMG, Paoletti FP, Gaetani L, Moda F, Parnetti L. α-Synuclein Seed Amplification Assays for Diagnosing Synucleinopathies: The Way Forward. Neurology. 2022;99(5):195–205. doi:10.1212/WNL.0000000000200878

81. Coysh T, Mead S. The Future of Seed Amplification Assays and Clinical Trials. Front Aging Neurosci. 2022;14:872629. doi:10.3389/FNAGI.2022.872629/BIBTEX

82. Bargar C, De Luca CMG, Devigili G, et al. Discrimination of MSA-P and MSA-C by RT-QuIC analysis of olfactory mucosa: the first assessment of assay reproducibility between two specialized laboratories. Mol Neurodegener. 2021;16(1). doi:10.1186/S13024-021-00491-Y

83. Parkkinen L, Soininen H, Laakso M, Alafuzoff I. Alpha-synuclein pathology is highly dependent on the case selection. Neuropathol Appl Neurobiol. 2001;27(4):314–325. doi:10.1046/J.0305-1846.2001.00342.X

